# Impact of Pulmonary and Sleep Disorders on COVID-19 Infection Severity in a Large Clinical Biobank

**DOI:** 10.1101/2023.02.19.23286148

**Authors:** Brian E. Cade, Syed Moin Hassan, Janet M. Mullington, Elizabeth W. Karlson, Susan Redline

**Affiliations:** Division of Sleep and Circadian Disorders, Brigham and Women’s Hospital, Boston, MA 02115, USA; Division of Sleep Medicine, Harvard Medical School, Boston, MA 02115, USA; Division of Pulmonary Disease and Critical Care Medicine, University of Vermont, Burlington, VT 05401, USA; Department of Neurology, Beth Israel Deaconess Medical Center, Boston, MA 02115, USA; Division of Rheumatology, Inflammation and Immunity, Brigham and Women’s Hospital, Boston, MA 02115, USA; Center for Genomic Medicine, Massachusetts General Hospital and Harvard Medical School, Boston, MA 02114, USA

**Keywords:** COVID-19, Epidemiology, Pulmonary disorders, Sleep disorders

## Abstract

**Rationale:** Multiple pulmonary, sleep, and other disorders are associated with the severity of Covid-19 infections but may or may not directly affect the etiology of acute Covid-19 infection. Identifying the relative importance of concurrent risk factors may prioritize respiratory disease outbreaks research.

**Objectives:** To identify associations of common preexisting pulmonary and sleep disease on acute Covid-19 infection severity, investigate the relative contributions of each disease and selected risk factors, identify sex-specific effects, and examine whether additional electronic health record (EHR) information would affect these associations.

**Methods:** 45 pulmonary and 6 sleep diseases were examined in 37,020 patients with Covid-19. We analyzed three outcomes: death; a composite measure of mechanical ventilation and/or ICU admission; and inpatient admission. The relative contribution of pre-infection covariates including other diseases, laboratory tests, clinical procedures, and clinical note terms was calculated using LASSO. Each pulmonary/sleep disease model was then further adjusted for covariates.

**Measurements and main results:** 37 pulmonary/sleep diseases were associated with at least one outcome at Bonferroni significance, 6 of which had increased relative risk in LASSO analyses. Multiple prospectively collected non-pulmonary/sleep diseases, EHR terms and laboratory results attenuated the associations between preexisting disease and Covid-19 infection severity. Adjustment for counts of prior “blood urea nitrogen” phrases in clinical notes attenuated the odds ratio point estimates of 12 pulmonary disease associations with death in women by ≥1.

**Conclusions:** Pulmonary diseases are commonly associated with Covid-19 infection severity. Associations are partially attenuated by prospectively-collected EHR data, which may aid in risk stratification and physiological studies.

## Introduction

Covid-19 continues to be an important public health problem. Prior to the Omicron wave, Covid-19 was one of the leading causes of death in the US in every age group above age 15 [1]. Thousands continue to die despite the availability of vaccines, emphasizing a continued need for research to help mitigate the effects of this and future infectious disease outbreaks.

The range of effects arising from Covid-19 infection, from minimal symptoms to death is striking. Less is known about the relative importance of individual pulmonary and sleep diseases contributing to infection severity in patients with multimorbidity. Examining the association of a range of pulmonary and sleep diseases with Covid-19 infection severity and applying machine learning methods to identify a subset of independent associations that predict infection severity may aid in identifying etiological mechanisms of morbidity and mortality. These associations may differ by sex, given sex differences in both Covid-19 prevalence and associated risk factors.

Associations between preexisting pulmonary/sleep disease and Covid-19 infection severity may be changing with time. The Omicron variant has reduced pulmonary pathology compared to other variants [2,3]. Patients carrying the Omicron variant have reduced risk of hospital admissions and severe infection [4]. The degree of association between individual diseases and Covid-19 severity may therefore change across different phases of the epidemic. Examining these changing environmental interactions may provide insight into the pathophysiology of both Covid-19 and the tested diseases themselves.

Prospective collection of data in clinical biobanks are one means of addressing these and other questions. EHR databases can leverage extensive clinical note, diagnosis, procedure, and laboratory data collected prior to infection. Natural language processing (NLP) [5] of controlled medical vocabulary concepts [6] from date-stamped clinical notes can improve disease phenotyping [7]. These clinical note concepts, together with procedure and laboratory information, can also be used to identify risk factors that complement or potentially mediate or confound the association of existing diseases with Covid-19 infection severity.

In this study, we report associations between common preexisting pulmonary and sleep diseases and acute Covid-19 morbidity and mortality among patients with PCR-based diagnoses of Covid-19 with data in the Mass General Brigham (MGB) EHR database from the greater Boston, MA region. We investigate the relative importance of individual preexisting diseases and ancillary EHR concepts, identify sex- and date-specific associations, and report the effect of lead EHR concepts on the associations of preexisting diseases and Covid-19 infection severity.

## Methods

Additional details are provided in the Supplemental Methods.

### Study sample

All patients with PCR-diagnosed Covid-19 were enrolled in the MGB Research Patient Data Registry EHR database [8,9]. Study collection ended at 6/1/2022. We used a stringent combined data floor and loyalty cohort [10] approach to minimize the number of patients with reduced documentation (and increased risk of false negative associations) [11] by requiring at least three clinical notes, encounters, and diagnoses. We also required at least 7 clinical facts: one diagnosis, encounter, laboratory result, medication, outpatient visit, routine visit [10], and vital sign obtained before 2020. Children, MGB employees, and patients living outside of Massachusetts or without available race/ethnicity, age, biological sex, or BMI values were excluded.

### Data used

The acute phase of infection for measuring ICU admissions, mechanical ventilation, and inpatient admissions was defined as -7 to +30 days relative to the first positive PCR test (we did not analyze Covid-19 reinfections) in the EHR. Diseases, procedures, and NLP terms recorded at least 8 days prior to the PCR test were extracted. Laboratory measurements were extracted from -5 to -1 years prior to the PCR test. A breakthrough infection was defined as having occurred over 30 days after a second vaccination. Majority variant date ranges were based on Broad Institute Covid-19 Genome Surveillance project sequencing in the greater Boston area. Transition dates from the pre-Delta to the Delta and Omicron variants were set at 7/3/2021 and 12/17/2021 respectively.

Diseases were based on PheCode groupings of ICD codes [12,13] (**Table S1**) and refined using natural language processing. We evaluated 45 common (≥1% prevalence in patients with Covid-19) and selected rare pulmonary disorders and 6 common sleep disorders [14–16]. Clinical note concept terms were extracted using NLP and mapped to Concept Unique Identifiers (CUIs) codes [5,6]. Terms that matched disease names were used to improve disease phenotyping using multimodal automated phenotyping (MAP) [7]. We included all non-pulmonary and non-sleep diseases defined using PheCodes and MAP with >1% prevalence among patients with Covid-19 in LASSO regression analyses (described below). Given the sparse nature of certain individual procedures, we also grouped procedures as specified by AHRQ Clinical Classification Software (CCS, https://www.hcup-us.ahrq.gov/toolssoftware/ccs10/ccs10.jsp). Procedure and CUI IDs for variables retained after LASSO regressions are listed in Table S6.

We documented evidence of continuous positive airway pressure (CPAP) use among patients with at least one clinical encounter and one clinical note from -365 to -8 days prior to the first positive PCR test. Total counts of five related procedures and one NLP term (Table S5) were combined. CPAP evidence in the prior year was present for 2,946 / 4,027 (73.1%) of the eligible patients with sleep apnea.

In an exploratory aim, we examined whether selected hematology laboratory results were associated with Covid-19 severity. We included kidney function measures given the retention of kidney disease in the LASSO results. The median values of laboratory results for individual patients from -5 to -1 years were tested with and without rank normalization. The 23 laboratory measurements were available for 65 – 89% of the patients (Table S8).

### Statistical analyses

Logistic regression included 45 pulmonary disorders, 6 sleep disorders, was adjusted for age, sex at birth, WHO obesity classification, race/ethnicity, and breakthrough infection status to predict 3 outcomes: death, mechanical ventilation or ICU admission, or inpatient admissions. The effect of CPAP evidence was tested using logistic regression with further adjustment for a single CPAP evidence term.

Adaptive LASSO regression used training and testing sets, each with cross-validation. Non-zero coefficient terms were carried forward to the training run and then to the final combined sample run. Age, sex, BMI, race/ethnicity, breakthrough infection status, healthcare encounter counts, and days into the pandemic were forced into runs. Laboratory results were not included due to reduced sample size and non-random ascertainment. We opted to include sex-specific terms in combined sex analyses and evaluate them in separate sex-stratified analyses. Finally, the potential effect of lead LASSO variables on disease associations with Covid-19 infection severity was assessed using logistic regression with standard covariates that included and excluded individual LASSO variables as covariates.

## Results

### Sample characteristics

Sample characteristics are listed in **Table 1**. The initial sample of patients with Covid-19 (through 6/1/2022) and suitable data to clear a data floor was 37,020. Women represented a majority of infections (62%) but a minority of patients with more severe death and mechanical ventilation, or ICU admission outcomes (48%). The majority of breakthrough infections were in women (62%). At the time of the data freeze (6/1/2022), the number of patients with an incident diagnosis during the pre-Delta majority variant virus dates was slightly less than the combined number of patients with a diagnosis during the Delta or Omicron majority variant diagnosis dates. The median BMI has decreased for Omicron majority variant diagnosis dates, as has the percentage of Hispanic/Latino Americans among patients with an incident Covid-19 infection. The prevalence of 45 pulmonary and 6 sleep diseases grouped by PheCodes (**Table S1**) may be reduced due to the strict case criteria used by NLP-defined phenotyping. The median number of preexisting pulmonary and sleep diseases diagnosed for patients with an adverse Covid-19 outcome was higher than the median number of preexisting diseases for all patients with Covid-19.

**Table 1.**
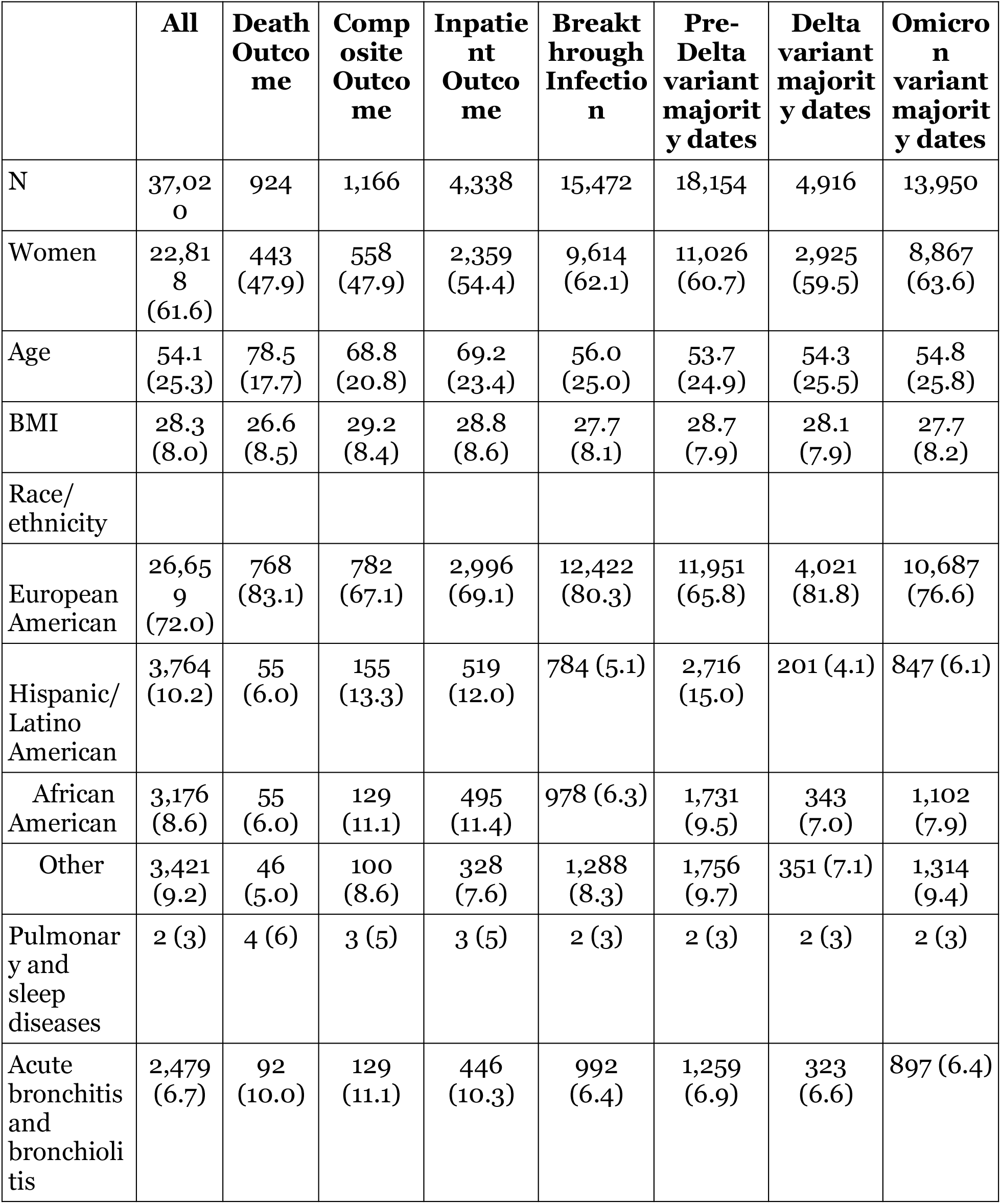
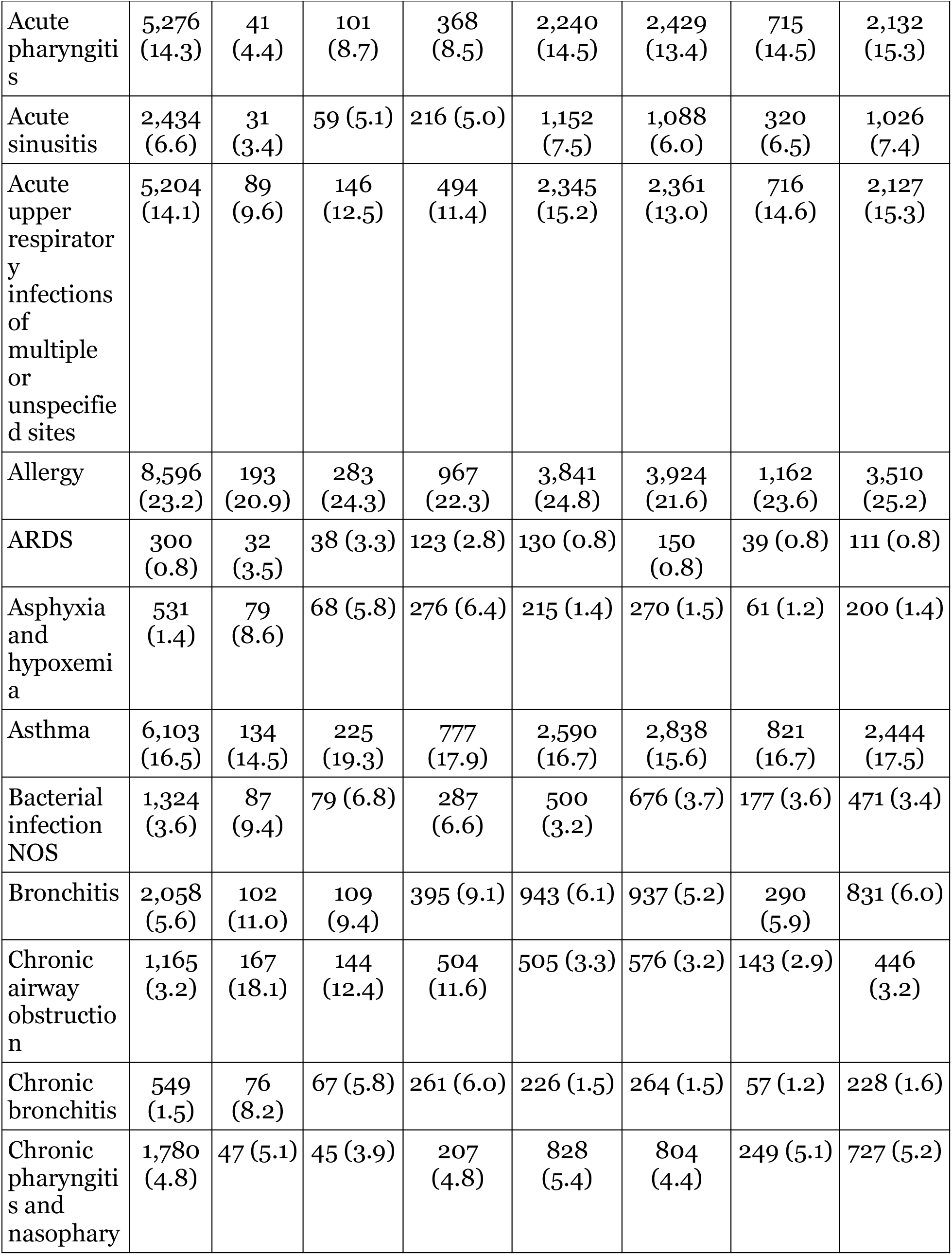
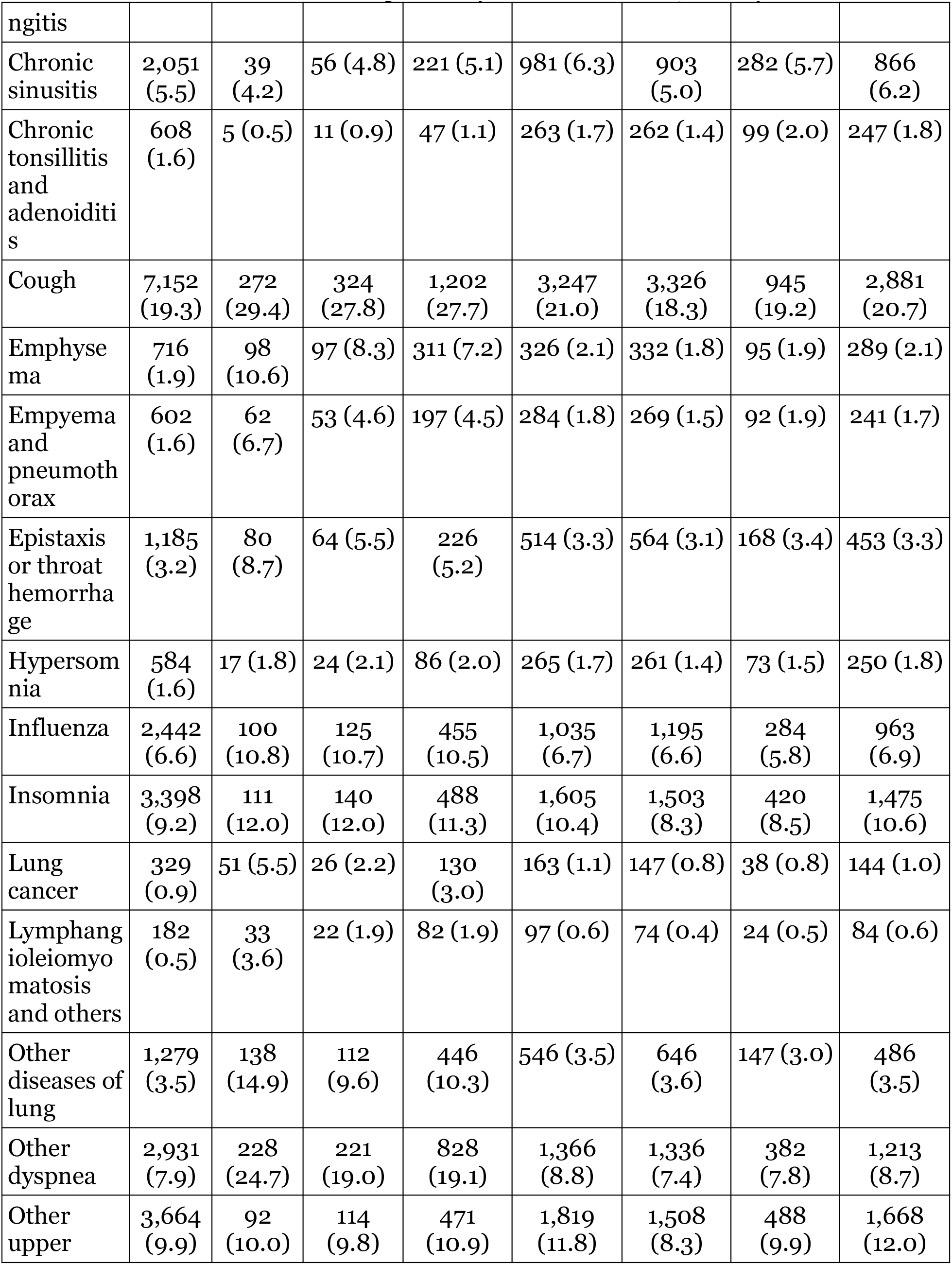
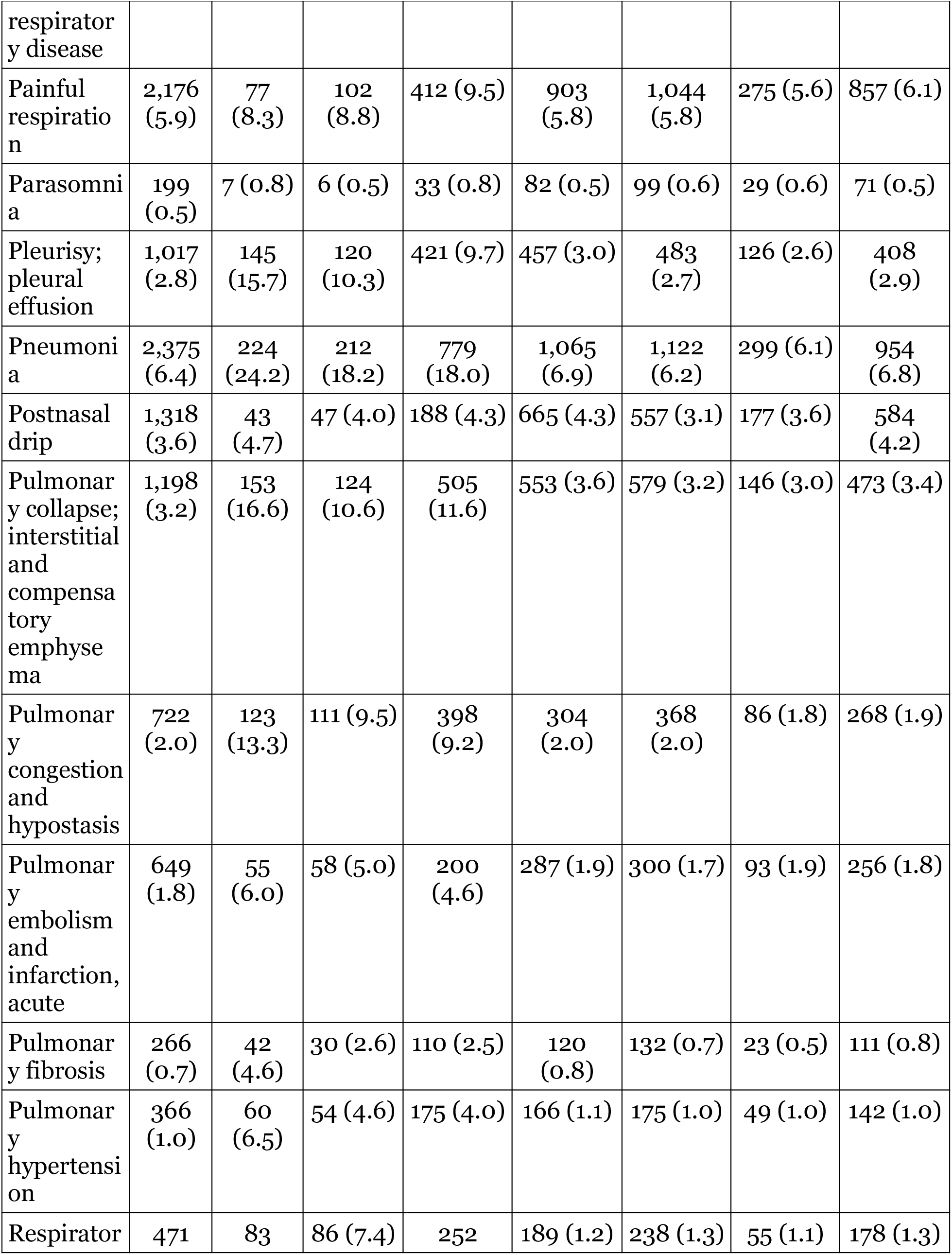
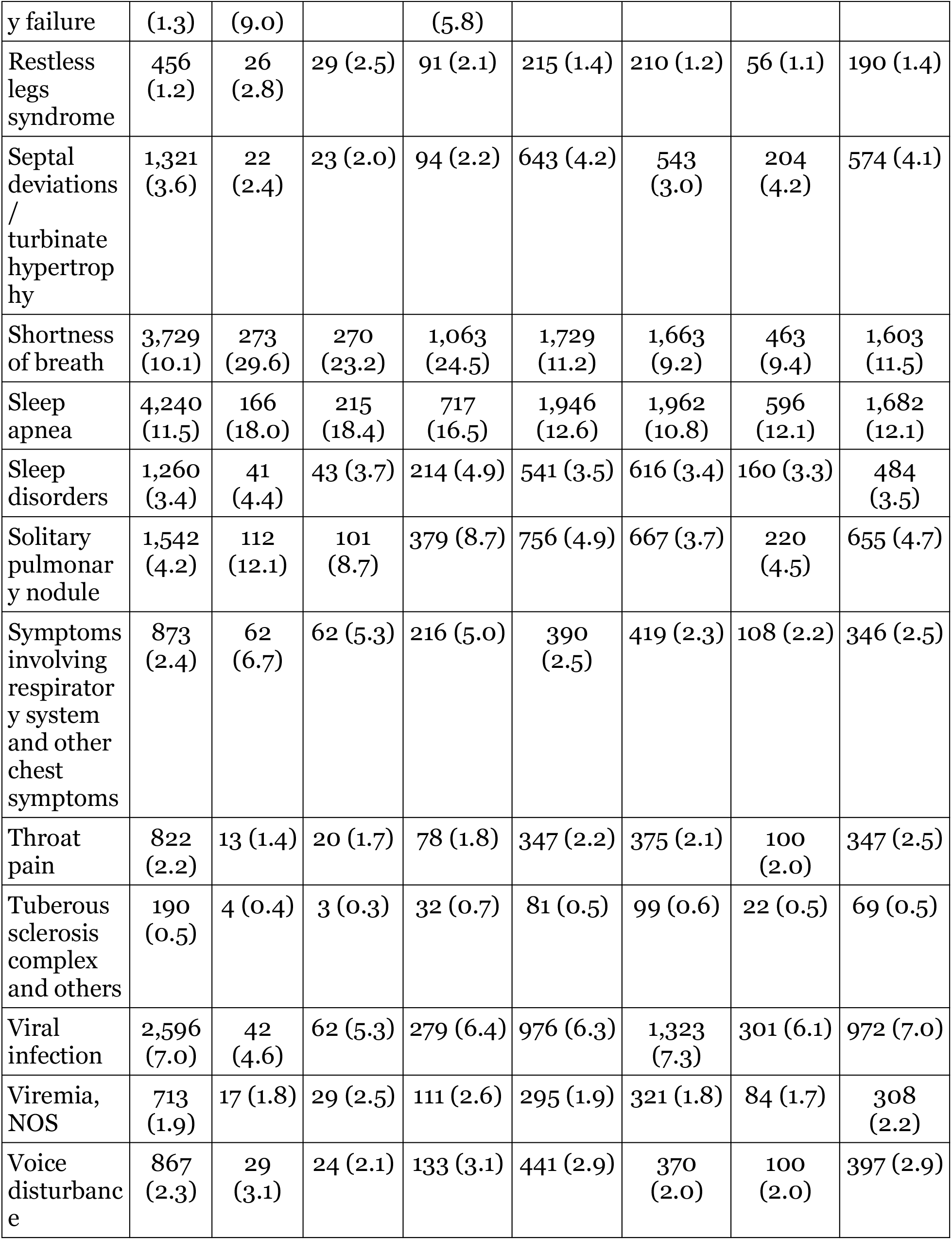
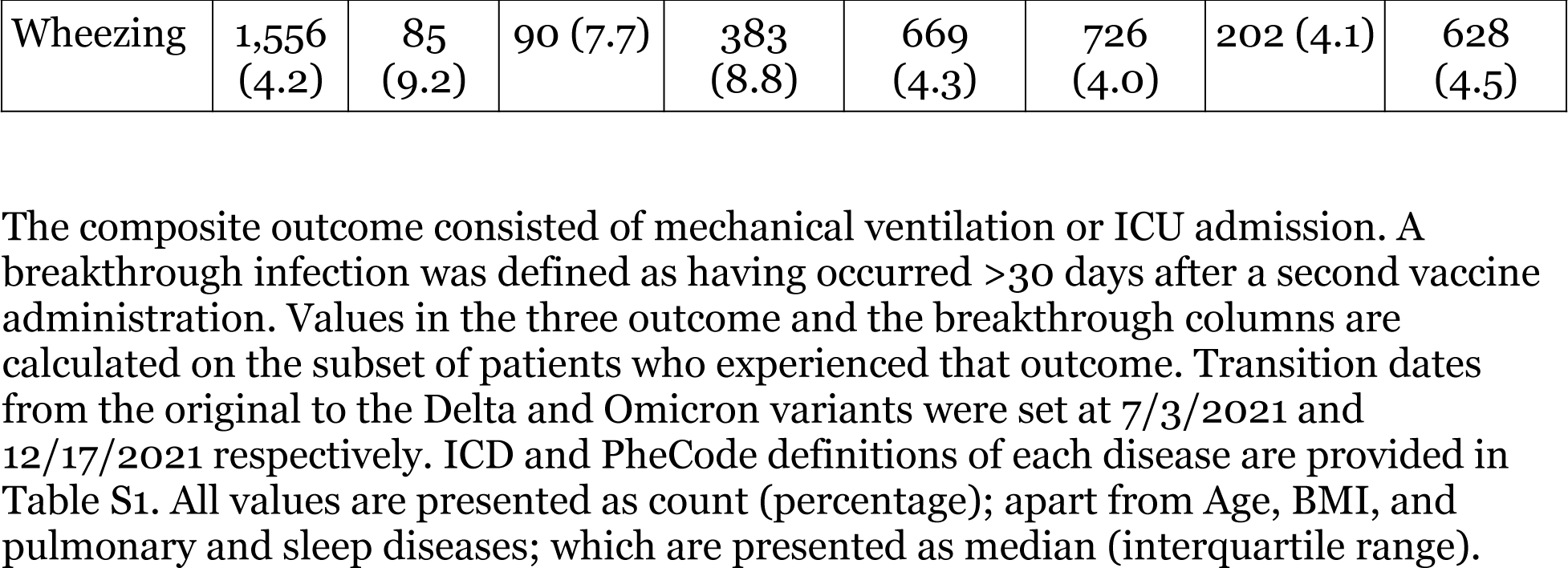
Sample characteristics.

### Associations between existing disease with Covid-19 infection severity

The associations between existing diseases and Covid-19 infection severity are described in **Table 2**. Models included adjustment for breakthrough infection, age, sex at birth, obesity severity, and self-reported race/ethnicity. 42 of 51 diseases were nominally associated with any outcome (p < 0.05). 37 diseases were associated with an outcome at Bonferroni significance (p < 3.3 × 10^−4^): 25, 23, and 37 diseases were associated with risk of death, mechanical ventilation or ICU admission, and inpatient admission, respectively. Chronic pharyngitis and nasopharyngitis and septal deviations / turbinate hypertrophy were inversely associated with risk of mechanical ventilation or ICU admission at modest significance.

**Table 2.** Associations between preexisting disease and Covid-19 infection severity.

| **Existing Disorder** | **Death Outcome** | **Mechanical ventilation or ICU admission Outcome** | **Inpatient admission Outcome** |
| --- | --- | --- | --- |
| Acute bronchitis and bronchiolitis | 1.25 (0.99 – 1.59) | **1.43 (1.18 – 1.74)<sup>†</sup>** | **1.42 (1.27 – 1.60)<sup>†</sup>** |
| Acute pharyngitis | 0.78 (0.56 – 1.08) | 1.05 (0.85 – 1.31) | 0.96 (0.85 – 1.08) |
| Acute sinusitis | 0.69 (0.48 – 1.01) | 0.94 (0.72 – 1.24) | 0.83 (0.72 – 0.97)<sup>*</sup> |
| Acute upper respiratory infections of multiple or unspecified sites | 0.88 (0.70 – 1.12) | 1.12 (0.93 – 1.35) | 0.92 (0.83 – 1.02) |
| Allergy | 1.13 (0.95 – 1.34) | 1.29 (1.12 – 1.49)<sup>*</sup> | 1.10 (1.01 – 1.19)<sup>*</sup> |
| ARDS | **2.48 (1.62 – 3.81)<sup>†</sup>** | **2.95 (2.03 – 4.29)<sup>†</sup>** | **3.90 (3.01 – 5.07)<sup>†</sup>** |
| Asphyxia and hypoxemia | **2.93 (2.21 – 3.89)<sup>†</sup>** | **2.36 (1.78 – 3.13)<sup>†</sup>** | **5.35 (4.41 – 6.49)<sup>†</sup>** |
| Asthma | 1.18 (0.97 – 1.44) | **1.40 (1.20 – 1.64)<sup>†</sup>** | **1.28 (1.17 – 1.40)<sup>†</sup>** |
| Bacterial infection NOS | **2.68 (2.06 – 3.48)<sup>†</sup>** | **1.96 (1.53 – 2.52)<sup>†</sup>** | **2.30 (1.98 – 2.68)<sup>†</sup>** |
| Bronchitis | 1.45 (1.16 – 1.83)<sup>*</sup> | 1.38 (1.12 – 1.70)<sup>*</sup> | **1.40 (1.24 – 1.59)<sup>†</sup>** |
| Chronic airway obstruction | **2.70 (2.22 – 3.28)<sup>†</sup>** | **2.42 (1.99 – 2.96)<sup>†</sup>** | **3.18 (2.79 – 3.63)<sup>†</sup>** |
| Chronic bronchitis | **2.30 (1.75 – 3.03)<sup>†</sup>** | **2.11 (1.59 – 2.78)<sup>†</sup>** | **3.53 (2.93 – 4.25)<sup>†</sup>** |
| Chronic pharyngitis and nasopharyngitis | 0.88 (0.64 – 1.21) | 0.72 (0.53 – 0.99)<sup>*</sup> | 0.89 (0.76 – 1.04) |
| Chronic sinusitis | 0.84 (0.60 – 1.18) | 0.97 (0.73 – 1.28) | 0.97 (0.84 – 1.13) |
| Chronic tonsillitis and adenoiditis | 1.06 (0.43 – 2.63) | 1.05 (0.57 – 1.94) | 1.23 (0.89 – 1.69) |
| Cough | 1.25 (1.07 – 1.46)<sup>*</sup> | **1.31 (1.14 – 1.51)<sup>†</sup>** | **1.39 (1.29 – 1.51)<sup>†</sup>** |
| Emphysema | **2.39 (1.87 – 3.05)<sup>†</sup>** | **2.90 (2.29 – 3.68)<sup>†</sup>** | **3.20 (2.71 – 3.77)<sup>†</sup>** |
| Empyema and pneumothorax | **2.38 (1.76 – 3.21)<sup>†</sup>** | **2.04 (1.50 – 2.76)<sup>†</sup>** | **2.65 (2.19 – 3.21)<sup>†</sup>** |
| Epistaxis or throat hemorrhage | **1.88 (1.45 – 2.45)<sup>†</sup>** | 1.33 (1.01 – 1.74)<sup>*</sup> | **1.35 (1.15 – 1.59)<sup>†</sup>** |
| Hypersomnia | 1.47 (0.88 – 2.46) | 1.27 (0.83 – 1.94) | 1.27 (1.00 – 1.63) |
| Influenza | 1.47 (1.16 – 1.85)<sup>*</sup> | **1.54 (1.26 – 1.88)<sup>†</sup>** | **1.68 (1.49 – 1.89)<sup>†</sup>** |
| Insomnia | 1.09 (0.88 – 1.35) | 1.26 (1.04 – 1.52)<sup>*</sup> | 1.09 (0.98 – 1.22) |

| Existing Disorder | Death Outcome | Mechanical ventilation or ICU admission Outcome | Inpatient admission Outcome |
| --- | --- | --- | --- |
| Lung cancer | **3.23 (2.31 – 4.52)<sup>†</sup>** | 1.55 (1.02 – 2.37)* | **2.87 (2.25 – 3.66)<sup>†</sup>** |
| Lymphangioleiomyomatosis | **4.57 (2.96 – 7.04)<sup>†</sup>** | **2.54 (1.57 – 4.11)<sup>†</sup>** | **3.82 (2.77 – 5.26)<sup>†</sup>** |
| Other diseases of lung | **2.13 (1.73 – 2.63)<sup>†</sup>** | **1.66 (1.34 – 2.05)<sup>†</sup>** | **2.33 (2.04 – 2.65)<sup>†</sup>** |
| Other dyspnea | **2.28 (1.92 – 2.70)<sup>†</sup>** | **1.85 (1.58 – 2.18)<sup>†</sup>** | **2.33 (2.12 – 2.57)<sup>†</sup>** |
| Other upper respiratory disease | 1.13 (0.89 – 1.42) | 1.09 (0.89 – 1.34) | **1.23 (1.10 – 1.37)<sup>†</sup>** |
| Painful respiration | 1.23 (0.96 – 1.60) | 1.33 (1.07 – 1.65)* | **1.58 (1.40 – 1.79)<sup>†</sup>** |
| Parasomnia | 1.36 (0.61 – 3.03) | 0.85 (0.37 – 1.96) | 1.39 (0.93 – 2.09) |
| Pleurisy; pleural effusion | **2.89 (2.33 – 3.58)<sup>†</sup>** | **2.45 (1.97 – 3.04)<sup>†</sup>** | **3.43 (2.97 – 3.96)<sup>†</sup>** |
| Pneumonia | **2.16 (1.81 – 2.58)<sup>†</sup>** | **2.13 (1.80 – 2.51)<sup>†</sup>** | **2.85 (2.57 – 3.16)<sup>†</sup>** |
| Postnasal drip | 0.89 (0.64 – 1.24) | 1.00 (0.73 – 1.35) | 1.02 (0.86 – 1.20) |
| Pulmonary collapse; interstitial and compensatory emphysema | **2.59 (2.10 – 3.18)<sup>†</sup>** | **2.05 (1.66 – 2.54)<sup>†</sup>** | **3.65 (3.19 – 4.17)<sup>†</sup>** |
| Pulmonary congestion and hypostasis | **3.55 (2.81 – 4.48)<sup>†</sup>** | **3.04 (2.41 – 3.82)<sup>†</sup>** | **5.85 (4.95 – 6.91)<sup>†</sup>** |
| Pulmonary embolism and infarction, acute | **1.87 (1.36 – 2.56)<sup>†</sup>** | **1.91 (1.42 – 2.56)<sup>†</sup>** | **2.26 (1.87 – 2.72)<sup>†</sup>** |
| Pulmonary fibrosis | **3.26 (2.22 – 4.78)<sup>†</sup>** | 2.09 (1.39 – 3.16)* | **3.03 (2.31 – 3.97)<sup>†</sup>** |
| Pulmonary hypertension | **3.15 (2.28 – 4.33)<sup>†</sup>** | **2.79 (2.03 – 3.83)<sup>†</sup>** | **3.67 (2.92 – 4.61)<sup>†</sup>** |
| Respiratory failure | **4.69 (3.54 – 6.21)<sup>†</sup>** | **4.44 (3.41 – 5.78)<sup>†</sup>** | **6.52 (5.31 – 8.01)<sup>†</sup>** |
| Restless legs syndrome | 1.58 (1.02 – 2.44)* | 1.61 (1.08 – 2.41)* | 1.38 (1.07 – 1.77)* |
| Septal deviations / turbinate hypertrophy | 0.85 (0.54 – 1.32) | 0.63 (0.41 – 0.96)* | **0.64 (0.52 – 0.80)<sup>†</sup>** |
| Shortness of breath | **2.04 (1.74 – 2.40)<sup>†</sup>** | **1.81 (1.56 – 2.10)<sup>†</sup>** | **2.48 (2.27 – 2.71)<sup>†</sup>** |
| Sleep apnea | **1.68 (1.38 – 2.03)<sup>†</sup>** | 1.32 (1.12 – 1.56)* | **1.27 (1.15 – 1.40)<sup>†</sup>** |
| Sleep disorders | 1.07 (0.76 – 1.50) | 0.88 (0.64 – 1.22) | **1.35 (1.15 – 1.59)<sup>†</sup>** |
| Solitary pulmonary | **1.64 (1.31 – 2.04)<sup>†</sup>** | 1.49 (1.20 – 1.86)* | **1.56 (1.37 – 1.77)<sup>†</sup>** |

**Cade *et al.*: Common pulmonary disease and Covid-19 severity**
| **Existing Disorder** | **Death Outcome** | **Mechanical ventilation or ICU admission Outcome** | **Inpatient admission Outcome** |
| --- | --- | --- | --- |
| nodule |  |  |  |
| Symptoms involving respiratory system and other chest symptoms | **2.27 (1.69 – 3.04)<sup>†</sup>** | **1.90 (1.44 – 2.51)<sup>†</sup>** | **1.98 (1.66 – 2.35)<sup>†</sup>** |
| Throat pain | 0.94 (0.53 – 1.67) | 0.92 (0.58 – 1.46) | 0.90 (0.70 – 1.16) |
| Tuberous Sclerosis Complex | 1.86 (0.66 – 5.28) | 0.71 (0.22 – 2.27) | **2.51 (1.66 – 3.80)<sup>†</sup>** |
| Viral infection | 1.47 (1.05 – 2.05)* | 1.16 (0.89 – 1.52) | **1.51 (1.31 – 1.74)<sup>†</sup>** |
| Viremia, NOS | 1.30 (0.77 – 2.19) | 1.52 (1.03 – 2.25)* | **1.71 (1.37 – 2.14)<sup>†</sup>** |
| Voice disturbance | 0.88 (0.59 – 1.31) | 0.71 (0.47 – 1.08) | 1.03 (0.85 – 1.26) |
| Wheezing | **1.76 (1.37 – 2.26)<sup>†</sup>** | 1.44 (1.14 – 1.81)* | **2.09 (1.83 – 2.38)<sup>†</sup>** |

Odds ratios with 95% confidence intervals are shown. All association calculations adjusted for age, sex, race/ethnicity, obesity category, and breakthrough infection status. \*: $p < 0.05$ , <sup>†</sup>: $p < 3.3 \times 10^{-4}$ (Bonferroni significance).

We analyzed sex-specific associations between existing diseases and Covid-19 infection severity. Death outcome results are shown in **Table 3**, with full results provided in **Table S2**. 27 and 33 diseases were associated with at least one infection severity outcome in men and women respectively at Bonferroni significance (p < 1.1 × 10^−4^). While multiple diseases displayed notably different odds ratio point estimates, sex × disease interaction p-values were generally modest in this cohort with stringent loyalty and data floor criteria. Associations between existing diseases and Covid-19 infection severity in different age groups are provided in **Table S3**.

**Table 3.** Sex-stratified associations between preexisting disease and death.

| Existing Disorder | Men | Women | P Sex Interaction |
| --- | --- | --- | --- |
| Acute bronchitis and bronchiolitis | 1.10 (0.77 – 1.59) | 1.35 (0.99 – 1.84) | 0.366 |
| Acute pharyngitis | 0.52 (0.28 – 0.97)* | 0.96 (0.65 – 1.42) | 0.104 |
| Acute sinusitis | 0.33 (0.14 – 0.77)* | 0.94 (0.61 – 1.43) | 0.035 |
| Acute upper respiratory infections of multiple or unspecified sites | 0.67 (0.45 – 1.00)* | 1.05 (0.78 – 1.40) | 0.063 |
| Allergy | 1.01 (0.77 – 1.32) | 1.23 (0.98 – 1.54) | 0.274 |
| ARDS | 2.76 (1.54 – 4.95)* | 2.15 (1.14 – 4.07)* | 0.621 |
| Asphyxia and hypoxemia | **2.68 (1.77 – 4.05)†** | **3.11 (2.11 – 4.57)†** | 0.486 |
| Asthma | 0.85 (0.61 – 1.19) | 1.42 (1.11 – 1.83)* | 0.012 |
| Bacterial infection NOS | **2.35 (1.57 – 3.53)†** | **2.93 (2.07 – 4.13)†** | 0.402 |
| Bronchitis | 1.22 (0.86 – 1.74) | 1.68 (1.25 – 2.27)* | 0.167 |
| Chronic airway obstruction | **2.56 (1.93 – 3.40)†** | **2.84 (2.16 – 3.72)†** | 0.667 |
| Chronic bronchitis | 1.89 (1.23 – 2.89)* | **2.64 (1.84 – 3.81)†** | 0.241 |
| Chronic pharyngitis and nasopharyngitis | 0.91 (0.59 – 1.41) | 0.86 (0.54 – 1.38) | 0.852 |
| Chronic sinusitis | 1.14 (0.73 – 1.77) | 0.60 (0.35 – 1.02) | 0.071 |
| Chronic tonsillitis and adenoiditis | 1.35 (0.41 – 4.42) | 0.82 (0.20 – 3.41) | 0.598 |
| Cough | 1.12 (0.89 – 1.40) | 1.37 (1.11 – 1.70)* | 0.109 |
| Emphysema | **2.32 (1.64 – 3.28)†** | **2.41 (1.70 – 3.42)†** | 0.976 |
| Empyema and pneumothorax | **2.68 (1.85 – 3.88)†** | 1.87 (1.10 – 3.19)* | 0.221 |
| Epistaxis or throat hemorrhage | 1.87 (1.30 – 2.69)* | 1.89 (1.29 – 2.76)* | 0.795 |
| Hypersomnia | 1.68 (0.90 – 3.13) | 1.21 (0.48 – 3.03) | 0.607 |
| Influenza | 1.41 (0.99 – 2.00) | 1.50 (1.10 – 2.06)* | 0.629 |

Cade *et al.*: Common pulmonary disease and Covid-19 severity
| Existing Disorder | Men | Women | P Sex Interaction |
| --- | --- | --- | --- |
| Insomnia | 1.25 (0.90 – 1.72) | 0.99 (0.74 – 1.33) | 0.353 |
| Lung cancer | 2.39 (1.44 – 3.97)* | **4.14 (2.63 – 6.51)<sup>†</sup>** | 0.110 |
| Lymphangioleiomyomatosis | **5.15 (2.93 – 9.04)<sup>†</sup>** | **4.07 (2.05 – 8.09)<sup>†</sup>** | 0.828 |
| Other diseases of lung | **1.86 (1.38 – 2.52)<sup>†</sup>** | **2.39 (1.79 – 3.18)<sup>†</sup>** | 0.210 |
| Other dyspnea | **2.10 (1.65 – 2.67)<sup>†</sup>** | **2.44 (1.91 – 3.10)<sup>†</sup>** | 0.203 |
| Other upper respiratory disease | 1.05 (0.75 – 1.46) | 1.22 (0.88 – 1.69) | 0.470 |
| Painful respiration | 1.37 (0.94 – 1.99) | 1.09 (0.76 – 1.55) | 0.381 |
| Parasomnia | 2.22 (0.88 – 5.58) | 0.42 (0.06 – 3.08) | 0.143 |
| Pleurisy; pleural effusion | **2.60 (1.94 – 3.47)<sup>†</sup>** | **3.27 (2.38 – 4.49)<sup>†</sup>** | 0.253 |
| Pneumonia | **1.98 (1.55 – 2.55)<sup>†</sup>** | **2.33 (1.82 – 3.00)<sup>†</sup>** | 0.387 |
| Postnasal drip | 1.08 (0.67 – 1.72) | 0.77 (0.48 – 1.24) | 0.399 |
| Pulmonary collapse; interstitial and compensatory emphysema | **2.61 (1.99 – 3.42)<sup>†</sup>** | **2.51 (1.81 – 3.46)<sup>†</sup>** | 0.935 |
| Pulmonary congestion and hypostasis | **4.31 (3.11 – 5.96)<sup>†</sup>** | **2.87 (2.05 – 4.02)<sup>†</sup>** | 0.170 |
| Pulmonary embolism and infarction, acute | 1.93 (1.24 – 3.00)* | 1.74 (1.11 – 2.73)* | 0.899 |
| Pulmonary fibrosis | **3.29 (1.90 – 5.71)<sup>†</sup>** | **3.35 (1.97 – 5.70)<sup>†</sup>** | 0.870 |
| Pulmonary hypertension | **3.62 (2.21 – 5.93)<sup>†</sup>** | **2.79 (1.84 – 4.25)<sup>†</sup>** | 0.570 |
| Respiratory failure | **4.12 (2.77 – 6.13)<sup>†</sup>** | **5.35 (3.58 – 7.99)<sup>†</sup>** | 0.364 |
| Restless legs syndrome | 2.08 (1.11 – 3.90)* | 1.28 (0.70 – 2.35) | 0.377 |
| Septal deviations / turbinate hypertrophy | 1.06 (0.63 – 1.79) | 0.53 (0.22 – 1.31) | 0.191 |

**Cade *et al.*: Common pulmonary disease and Covid-19 severity**
| **Existing Disorder** | **Men** | **Women** | **P Sex Interaction** |
| --- | --- | --- | --- |
| Shortness of breath | **1.84 (1.46 – 2.32)<sup>†</sup>** | **2.22 (1.78 – 2.78)<sup>†</sup>** | 0.123 |
| Sleep apnea | **1.68 (1.31 – 2.15)<sup>†</sup>** | 1.79 (1.32 – 2.44)* | 0.306 |
| Sleep disorders | 1.08 (0.66 – 1.75) | 1.07 (0.66 – 1.74) | 0.914 |
| Solitary pulmonary nodule | 1.56 (1.14 – 2.14)* | 1.65 (1.21 – 2.25)* | 0.754 |
| Symptoms involving respiratory system and other chest symptoms | 2.13 (1.38 – 3.30)* | **2.37 (1.58 – 3.53)<sup>†</sup>** | 0.711 |
| Throat pain | 0.56 (0.17 – 1.80) | 1.20 (0.61 – 2.33) | 0.244 |
| Tuberous Sclerosis Complex | 0.00 (0.00 – Inf) | 3.13 (1.06 – 9.28)* | 0.945 |
| Viral infection | 1.36 (0.82 – 2.25) | 1.54 (0.99 – 2.42) | 0.746 |
| Viremia, NOS | 0.71 (0.28 – 1.84) | 1.85 (0.99 – 3.46) | 0.093 |
| Voice disturbance | 0.96 (0.54 – 1.70) | 0.82 (0.48 – 1.43) | 0.747 |
| Wheezing | **2.03 (1.41 – 2.93)<sup>†</sup>** | 1.53 (1.08 – 2.17)* | 0.441 |

We examined associations between existing disease and Covid-19 incident infection severity during days when the pre-Delta, Delta, and Omicron virus variants comprised the majority of samples sequenced from the greater Boston area. **Table 4** lists the composite outcome results, with full results and sex-stratified results listed in **Table S4**. The pre-Delta, Delta, and Omicron majority variant date analyses were all adjusted for breakthrough infection status. 31 and 26 diseases were associated with any outcome during pre-Delta and Omicron variant majority dates respectively (Bonferroni p < 1.1 × 10^−4^). Seven diseases significantly associated with a death outcome during pre-Delta variant majority dates (p < 1.1 × 10^−4^) were notably attenuated (p > 0.01) during Omicron variant majority dates (bacterial infection NOS, empyema and pneumothorax, epistaxis or throat hemorrhage, pneumonia, pulmonary hypertension, sleep apnea, and symptoms involving respiratory system and other chest symptoms). Conversely, a modest association between solitary pulmonary nodule and the death outcome during pre-Delta variant majority dates strengthened to Bonferroni significance during Omicron variant majority dates.

**Table 4.** Date-specific associations between preexisting disease and death.

| Existing disease | Pre-Delta variant majority dates | Delta variant majority dates | Omicron variant majority dates |
| --- | --- | --- | --- |
| Acute bronchitis and bronchiolitis | 1.25 (0.94 – 1.67) | 1.08 (0.52 – 2.25) | 1.25 (0.75 – 2.08) |
| Acute pharyngitis | 0.79 (0.52 – 1.19) | 0.53 (0.16 – 1.73) | 0.91 (0.50 – 1.67) |
| Acute sinusitis | 0.65 (0.40 – 1.06) | 0.65 (0.23 – 1.82) | 0.81 (0.39 – 1.68) |
| Acute upper respiratory infections of multiple or unspecified sites | 0.89 (0.66 – 1.20) | 1.31 (0.73 – 2.35) | 0.71 (0.43 – 1.16) |
| Allergy | 1.12 (0.90 – 1.40) | 1.02 (0.60 – 1.71) | 1.20 (0.85 – 1.70) |
| ARDS | 2.55 (1.47 – 4.40)* | 2.77 (0.88 – 8.71) | 2.33 (0.96 – 5.63) |
| Asphyxia and hypoxemia | **3.39 (2.40 – 4.80)<sup>†</sup>** | 1.53 (0.50 – 4.65) | 2.52 (1.40 – 4.53)* |
| Asthma | 1.13 (0.88 – 1.46) | 1.24 (0.69 – 2.22) | 1.34 (0.91 – 1.97) |
| Bacterial infection NOS | **3.00 (2.16 – 4.16)<sup>†</sup>** | 3.91 (1.89 – 8.05)* | 1.63 (0.90 – 2.95) |
| Bronchitis | 1.45 (1.09 – 1.94)* | 1.43 (0.73 – 2.80) | 1.43 (0.90 – 2.26) |
| Chronic airway obstruction | **2.59 (2.02 – 3.33)<sup>†</sup>** | 2.19 (1.15 – 4.18)* | **3.47 (2.39 – 5.04)<sup>†</sup>** |
| Chronic bronchitis | **2.26 (1.58 – 3.23)<sup>†</sup>** | 1.84 (0.69 – 4.91) | **3.07 (1.87 – 5.03)<sup>†</sup>** |
| Chronic pharyngitis and nasopharyngitis | 0.86 (0.57 – 1.30) | 1.25 (0.58 – 2.71) | 0.79 (0.40 – 1.56) |
| Chronic sinusitis | 0.90 (0.59 – 1.39) | 0.99 (0.42 – 2.35) | 0.64 (0.30 – 1.35) |
| Chronic tonsillitis and adenoiditis | 1.90 (0.74 – 4.84) | N/A | N/A |
| Cough | 1.15 (0.94 – 1.40) | 1.71 (1.09 – 2.69)* | 1.30 (0.95 – 1.78) |
| Emphysema | **2.35 (1.71 – 3.24)<sup>†</sup>** | 1.91 (0.87 – 4.22) | **2.86 (1.83 – 4.48)<sup>†</sup>** |
| Empyema and pneumothorax | **2.83 (1.95 – 4.12)<sup>†</sup>** | 1.24 (0.45 – 3.42) | 1.94 (1.03 – 3.65)* |
| Epistaxis or throat hemorrhage | **1.97 (1.42 – 2.74)<sup>†</sup>** | 1.63 (0.71 – 3.72) | 1.69 (0.99 – 2.87) |

Cade *et al.*: Common pulmonary disease and Covid-19 severity
| Existing disease | Pre-Delta variant majority dates | Delta variant majority dates | Omicron variant majority dates |
| --- | --- | --- | --- |
| Hypersomnia | 1.23 (0.61 – 2.50) | 1.23 (0.25 – 6.10) | 2.30 (0.98 – 5.40) |
| Influenza | 1.72 (1.29 – 2.30)* | 1.57 (0.79 – 3.13) | 0.90 (0.53 – 1.55) |
| Insomnia | 1.17 (0.89 – 1.54) | 1.61 (0.89 – 2.92) | 0.78 (0.49 – 1.22) |
| Lung cancer | **3.44 (2.20 – 5.38)<sup>†</sup>** | 1.52 (0.43 – 5.45) | **3.75 (2.11 – 6.65)<sup>†</sup>** |
| Lymphangioliomyomatosis | **4.76 (2.59 – 8.72)<sup>†</sup>** | 2.08 (0.45 – 9.72) | **5.42 (2.69 – 10.93)<sup>†</sup>** |
| Other diseases of lung | **2.12 (1.63 – 2.75)<sup>†</sup>** | 1.27 (0.61 – 2.65) | **2.69 (1.81 – 3.98)<sup>†</sup>** |
| Other dyspnea | **2.20 (1.76 – 2.74)<sup>†</sup>** | **2.79 (1.71 – 4.54)<sup>†</sup>** | **2.26 (1.61 – 3.17)<sup>†</sup>** |
| Other upper respiratory disease | 1.23 (0.92 – 1.66) | 1.15 (0.59 – 2.24) | 0.98 (0.62 – 1.56) |
| Painful respiration | 1.19 (0.86 – 1.65) | 0.69 (0.27 – 1.77) | 1.64 (1.02 – 2.66)* |
| Parasomnia | 1.90 (0.77 – 4.71) | N/A | 0.81 (0.11 – 6.14) |
| Pleurisy; pleural effusion | **2.89 (2.18 – 3.81)<sup>†</sup>** | **4.44 (2.37 – 8.31)<sup>†</sup>** | **2.61 (1.73 – 3.96)<sup>†</sup>** |
| Pneumonia | **2.38 (1.91 – 2.97)<sup>†</sup>** | 2.50 (1.47 – 4.26)* | 1.59 (1.10 – 2.30)* |
| Postnasal drip | 0.85 (0.54 – 1.32) | 1.45 (0.64 – 3.30) | 0.81 (0.42 – 1.54) |
| Pulmonary collapse; interstitial and compensatory emphysema | **2.33 (1.78 – 3.05)<sup>†</sup>** | 3.13 (1.70 – 5.78)* | **3.00 (2.03 – 4.42)<sup>†</sup>** |
| Pulmonary congestion and hypostasis | **3.43 (2.54 – 4.63)<sup>†</sup>** | **5.37 (2.80 – 10.29)<sup>†</sup>** | **3.06 (1.90 – 4.94)<sup>†</sup>** |
| Pulmonary embolism and infarction, acute | 1.68 (1.12 – 2.53)* | 1.17 (0.44 – 3.12) | 2.92 (1.66 – 5.16)* |
| Pulmonary fibrosis | **3.63 (2.25 – 5.86)<sup>†</sup>** | 1.33 (0.16 – 10.78) | 2.96 (1.41 – 6.20)* |
| Pulmonary hypertension | **3.25 (2.17 – 4.88)<sup>†</sup>** | 3.82 (1.63 – 8.98)* | 2.33 (1.16 – 4.69)* |
| Respiratory failure | **5.62 (3.92 – 8.07)<sup>†</sup>** | **5.36 (2.34 – 12.27)<sup>†</sup>** | **3.51 (1.99 – 6.17)<sup>†</sup>** |

Cade *et al.*: Common pulmonary disease and Covid-19 severity
| Existing disease | Pre-Delta variant majority dates | Delta variant majority dates | Omicron variant majority dates |
| --- | --- | --- | --- |
| Restless legs syndrome | 1.65 (0.96 – 2.84) | 0.71 (0.09 – 5.41) | 1.83 (0.83 – 4.07) |
| Septal deviations / turbinate hypertrophy | 1.22 (0.72 – 2.05) | 0.57 (0.14 – 2.40) | 0.42 (0.13 – 1.33) |
| Shortness of breath | **2.09 (1.71 – 2.57)<sup>†</sup>** | 1.75 (1.06 – 2.87)* | **2.16 (1.58 – 2.96)<sup>†</sup>** |
| Sleep apnea | **1.83 (1.44 – 2.33)<sup>†</sup>** | 1.64 (0.94 – 2.88) | 1.35 (0.89 – 2.04) |
| Sleep disorders | 1.39 (0.94 – 2.06) | 0.48 (0.11 – 2.03) | 0.57 (0.23 – 1.41) |
| Solitary pulmonary nodule | 1.47 (1.09 – 1.97)* | 1.37 (0.71 – 2.63) | **2.24 (1.52 – 3.29)<sup>†</sup>** |
| Symptoms involving respiratory system and other chest symptoms | **2.31 (1.57 – 3.39)<sup>†</sup>** | 2.54 (1.16 – 5.57)* | 2.07 (1.14 – 3.76)* |
| Throat pain | 0.78 (0.33 – 1.82) | 1.44 (0.33 – 6.31) | 1.19 (0.47 – 3.03) |
| Tuberous Sclerosis Complex | 1.85 (0.55 – 6.25) | N/A | 1.92 (0.23 – 15.79) |
| Viral infection | 1.44 (0.94 – 2.20) | 1.24 (0.38 – 4.12) | 1.83 (0.99 – 3.39) |
| Viremia, NOS | 1.77 (0.97 – 3.23) | N/A | 0.91 (0.28 – 2.96) |
| Voice disturbance | 0.98 (0.59 – 1.61) | 0.24 (0.03 – 1.81) | 0.98 (0.48 – 1.99) |
| Wheezing | 1.83 (1.33 – 2.51)* | 2.19 (1.08 – 4.46)* | 1.60 (0.95 – 2.70) |

### Effects of continuous airway positive pressure

We examined whether adjusting for evidence of continuous positive airway pressure (CPAP) usage in clinical notes and procedures in the year prior to infection would attenuate the associations with sleep apnea, given the trends we identified in our previous report with a smaller sample size [14]. All associations with sleep apnea were attenuated (p > 0.02, **Table S5**).

### Relative importance of individual existing diseases and EHR information

Given the large number of associated diseases and elevated multimorbidity (18.8% of the patients carried ≥5 of the tested diseases), we analyzed the relative importance of individual existing diseases affecting the risk of Covid-19 infection severity. We performed adaptive LASSO in the combined sex and sex-stratified datasets by including all pulmonary and sleep diseases and multiple covariates. Non-zero coefficients from the outcome results are shown in **Tables 5 and S6**, with larger absolute values indicating greater importance for individual variables. Chronic airway obstruction, pneumonia, and respiratory failure had non-zero coefficients for the death outcome.

**Table 5.** Joint pulmonary and sleep disorders LASSO regression nonzero coefficients.

| **Disease or covariate** | **Death Outcome** | **Mechanical ventilation or ICU admission Outcome** | **Inpatient admission Outcome** |
| --- | --- | --- | --- |
| Age | 1.610 | 0.780 | 0.826 |
| BMI | 0 | 0.076 | 0 |
| Sex (1M, 2F) | -0.089 | -0.149 | -0.042 |
| Breakthrough Infection | 0 | -0.074 | -0.250 |
| Encounters | 0.071 | 0.116 | 0.296 |
| Days into pandemic (1/1/2020) | -0.442 | -0.734 | -0.222 |
| Emphysema | N/A | 0.071 | N/A |
| Pneumonia | 0.039 | N/A | 0.105 |
| Chronic airway obstruction | 0.093 | N/A | N/A |
| Pulmonary congestion and hypostasis | N/A | 0.059 | 0.097 |
| Respiratory failure | 0.091 | N/A | 0.075 |
| Shortness of breath | N/A | N/A | 0.141 |

We next leveraged additional EHR information by performing similar adaptive LASSO analyses to identify factors with greater importance using separate analyses of 505 non-pulmonary/sleep diseases, 1,647 procedures, 171 CCS grouped procedures, and 9,839 clinical note NLP terms. Results for non-zero coefficient terms are listed in **Table S6**. 124 EHR variables were identified in the combined sex analyses. Six non-pulmonary/sleep diseases and ICD-coded procedures had non-zero coefficient terms in the combined-sex death outcome analysis: chronic renal failure, chemotherapy, altered mental status, essential hypertension, thrombocytopenia, and abnormality of gait. Associations between LASSO terms and Covid-19 infection severity adjusted for covariates are presented in **Table S7**. Multiple non-pulmonary diseases were associated with the death outcome, including chronic renal failure (OR 3.61, 95% CI 3.04 – 4.28) and thrombocytopenia (OR 3.19, 95% CI 2.45 – 4.17).

### Predictive laboratory values

Complete blood count and other hematology measurements are commonly collected in many patients undergoing hospital procedures. As an exploratory aim, we examined whether the median values of these measurements collected 1 – 5 years prior to the incident Covid-19 diagnosis were associated with infection severity. We also included measures of kidney function, given the identification of kidney disease in the LASSO results. 17 of the 23 laboratory tests were associated with at least one outcome in the available sample at Bonferroni significance (65 – 89% coverage of patients), using logistic regression with adjustment for covariates (**Table S8**). Estimated glomerular filtration rate, hematocrit, hemoglobin, and red cell distribution width results were the most notable associated measures with all p-values ≤ 2.0 × 10^−16^, despite the identification of thrombocytopenia and elevated white blood cell counts PheCodes in the LASSO results.

### Associated EHR concepts attenuate disease associations

Finally, we examined whether individual retained LASSO terms and prior hematology and kidney laboratory results would attenuate the associations between individual diseases and infection severity when included individually as model covariate terms. The lead terms with the highest median point estimate adjustment differences across all disease associations with the death outcome are shown in **Figure 1**. Lead terms spanned a range of healthcare procedures, laboratory results, and pulmonary and non-pulmonary diseases. Full results are provided in **Table S9**, and additional visualizations are provided in **Figures S1 – S17**. Adjustment for “blood urea nitrogen” clinical note phases attenuated the odds ratio point estimates of 12 pulmonary disease associations with death in women by ≥ 1.0.

**Figure 1.**
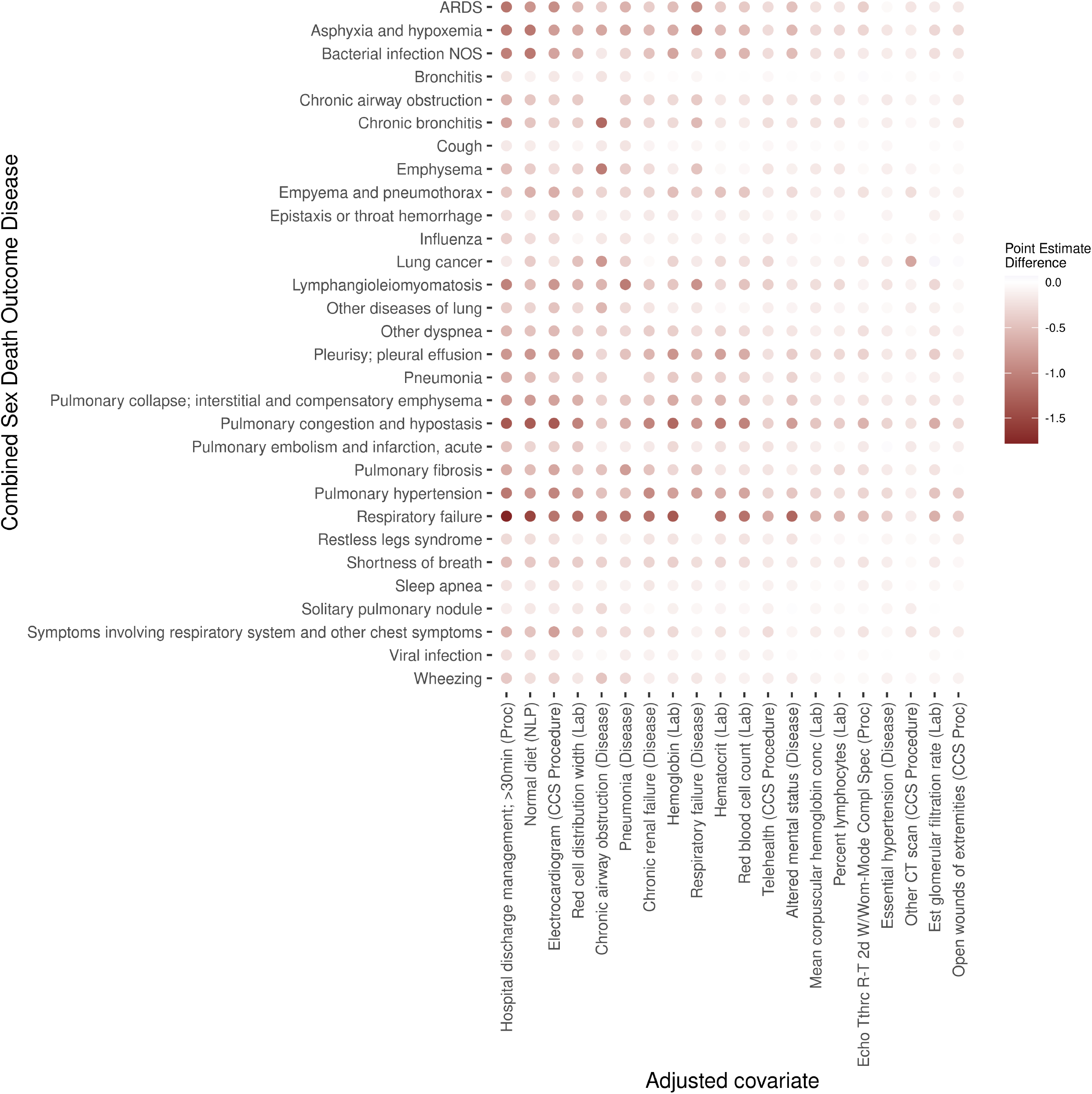
Effect of lead LASSO terms and laboratory results on disease associations with Covid-19 death outcome. The y axis includes all pulmonary and sleep diseases associated with death in the combined sex analyses (Table 2). Lead LASSO terms (Table S6) and laboratory results (Table S8) were included individually as covariates in each of these disease associations. The odds ratio difference when including versus not including an adjusted covariate are shown as circles, with darker red indicating an attenuated odds ratio for the Y-axis disease association with death when adjusting for the X-axis covariate. The X-axis covariates are ordered by median value of point estimates considering all of the Y-axis diseases. Additional visualizations considering the three outcomes presented as point estimate and p-value differences are provided in Figures S1 – S15. Full results with sortable values are provided in Table S9.

## Discussion

In this study, we examined the association of preexisting pulmonary, sleep, and other diseases with the risk of acute Covid-19 infection severity and investigated other factors that may alter those associations. Notable results include (a) common associations between dozens of pulmonary disorders and infection severity, some of which are protective; (b) changes in disease associations across date periods with different SARS-CoV-2 majority variants, with attenuated associations with death for 7 diseases, and an increased point estimate for 1 disease; (c) an increased importance of specific pulmonary disease associations with Covid-19 severity relative to other comorbid pulmonary and sleep diseases; (d) associations between infection severity and multiple hematology and kidney-related laboratory results collected 1 – 5 years prior to infection; and (e) non-pulmonary/sleep diseases, laboratory results, hospital procedures, and clinical note terms that attenuate and sometimes increase the associations between pulmonary and sleep diseases with Covid-19 infection severity. These results may aid in further understanding the etiology of a changing disease.

We observed common associations between preexisting pulmonary and sleep diseases with Covid-19 infection severity, including 37 of the 51 tested diseases associated with at least one outcome at Bonferroni significance (Tables 2 and S2). Certain diseases (e.g. asthma) can confer different risks for men and women, although sex × disease interaction p-values tended to be modest (Tables 3 and S2). Acute sinusitis has a protective association with the death measure in men (0.33 OR, 95% CI 0.14 – 0.77, p = 9.8 × 10^−3^) that is not present in women (0.94 OR, 95% CI 0.61 – 1.43, p = 0.76). A similar but more modest association was also observed between the death outcome and acute upper respiratory infections. These findings may help improve the personalization of treatment for patients with increased Covid-19 morbidity.

The associations between preexisting disease and infection severity are changing across time for certain diseases (Tables 4 and S4). Notable diseases with attenuated associations with death during Omicron-majority variant periods compared to pre-Delta majority variant periods include bacterial infection NOS (3.00 OR [95% CI 2.16 - 4.16] pre-Delta period; 1.63 [0.90 - 2.95] Omicron period), sleep apnea (1.83 [1.44 - 2.33] pre-Delta period; 1.35 [0.89 - 2.04] Omicron period), and influenza (1.72 [1.29 - 2.30] pre-Delta period; 0.90 [0.53 - 1.55] Omicron period). These changes may reflect differences in SARS-CoV-2 pathogenicity in upper and lower respiratory tracts [2,3]. The differences in outcomes across majority variant strain periods are consistent with a recent report examining the same dataset using independent methods [17]. Differences in risk factors and outcomes have potential study design implications for acute and possibly Long Covid research. The changing nature of SARS-CoV-2 associations with a range of comorbidities could potentially provide a window for future physiological research examining how individual pulmonary and sleep diseases increase risk for outcomes in other diseases.

Our study sample has a high degree of multimorbidity, and 39 pre-existing diseases were associated with at least one infection severity outcome at Bonferroni significance. We therefore performed LASSO regression analyses to identify the relative importance of specific diseases (Tables 5 and S6). LASSO analyses minimize the number of factors that collectively may predict an outcome. Six pulmonary diseases were retained: chronic airway obstruction, respiratory failure, pneumonia, emphysema, pulmonary congestion and hypostasis, and shortness of breath (Table 5). These diseases may be enriched with pathophysiological mechanisms that contribute increased risk across the broader range of pulmonary and sleep disorders associated with acute Covid-19 infection severity.

Preexisting non-pulmonary diseases, hospital procedures and clinical note terms identified by LASSO regression and laboratory results may have predictive value for Covid-19 infection severity, given the common associations between preexisting pulmonary disease and Covid-19 infection severity (Table S6). These variables may potentially mediate or confound associations between preexisting pulmonary and sleep diseases and infection severity (Table S9 and Figures 1, S1 – S17) or could reflect the effects of other factors unmeasured in this study. Five non-pulmonary PheCode-encoded diseases (and chemotherapy) were associated with death: chronic renal failure, altered mental status, essential hypertension, thrombocytopenia, and abnormality of gait. We chose to analyze procedures and all NLP term with common frequencies in order to identify factors that could help predict outcome severity, even if they are unlikely to be directly related to pathophysiology. Some lead terms that attenuate the associations of multiple diseases are related to healthcare utilization (e.g. hospital discharge procedures and “normal diet” text phrases) that are consistent with multimorbidity increasing the risk of adverse Covid-19 infection outcomes. Adjustment for electrocardiogram procedures and red cell distribution width laboratory results also attenuated the median point estimate of pulmonary and sleep disease associations with the death outcome more than adjustment for any pulmonary disease. The most notable variable to broadly affect associations with death was “blood urea nitrogen” in analyses of women (Figures S12 and S13), which attenuated the point estimates of 12 diseases by >1 and the statistical significance of 13 diseases (p > 0.05), although a smaller effect was observed when adjusting for actual median urea nitrogen blood levels collected from -5 to -1 years prior to infection with Covid-19. Adjustment for chronic renal failure and creatinine and estimated glomerular filtration rate laboratory results also support a role for kidney malfunction contributing to pulmonary and sleep disease associations with deaths in women.

Seventeen of 23 prospectively collected hematology and kidney function laboratory tests were associated with Covid-19 outcome severity at Bonferroni significance (Table S8). Estimated glomerular filtration rate and three red blood cell tests were the most significantly associated tests across all models. The laboratory value ranges may reflect physiological changes due to both known and unknown risk factors and may be useful as a general risk stratification measure. These associations were based on a subset of patients with available data, and the associations could possibly also reflect differences in test ordering (e.g. kidney test results may be disproportionally missing for patients whose clinicians do not suspect kidney malfunction). Additional follow-up in a broader range of patients is needed to understand the generalization of these results. The prediction models could be readily improved beyond these initial exploratory results.

The strengths of this study include an extensive sample size of patients diagnosed using PCR testing, which has allowed us to perform sex-, age-, and majority SARS-CoV-2 strain date-specific analyses with increased study power. Data were restricted by stringent combined “data floor” and loyalty cohort criteria. We included patients with minimal clinical encounters, notes, diagnoses and evidence of seven clinical facts each collected before the pandemic in order to increase the reliability of our findings in an open environment where some patient records may be located in other healthcare systems. All disease and healthcare utilization information was collected prior to Covid-19 infection to eliminate potential reverse causation effects. All disease phenotyping included natural language processing to increase the positive predictive value in the face of rule-out diagnoses and other potential biases. We extensively studied the potential effect of dozens of pulmonary disease on Covid-19 infection severity, which enabled us to identify the relative importance of pulmonary and disease on infection severity.

Weaknesses are also possible with this study design. Despite inclusion of over 37,000 patients, this was a single healthcare system study. While the Mass General Brigham system is comprised of multiple hospitals, our results may or may not completely generalize to other healthcare systems. Loss to follow-up and missing data from out-of-network care may affect our results and those of similarly designed studies. Temporal effects may be confounded by changes in PCR testing availability and use and standards of care (e.g. proning). Patients with different financial means may have different levels of healthcare utilization, which may influence the recording of diseases, procedures, and NLP phrases differently. The diseases, procedures, and note terms identified by LASSO regression may not completely generalize, despite cross-validation and separate training and testing sets. Individual terms with lower LASSO coefficients, terms that were not individually associated with outcomes (Table S7), or terms that do not affect multiple disease associations may have increased likelihood of site-specific associations or may indirectly measure factors not included in our study design. While the majority of the patients had undergone all of the laboratory testing (Table S8), there is likely some selection bias present for patients with clinically relevant diagnoses, although some of our results are supported by orthogonal diagnosis and NLP term data (Table S7).

## Conclusions

We identified common associations between preexisting pulmonary disease and Covid-19 infection severity and identified specific diseases with increased relative importance in our patient sample with multimorbidity. Certain associations are majority-variant date specific. We also identified several non-pulmonary diseases, clinical procedures, and clinical note phrases that potentially mediate or confound these associations. These results may aid in further understanding the etiology of a changing disease.

## Data Availability

All data produced in the present work are contained in the manuscript

## Acknowledgments

The authors wish to thank the MGB Research Patient Data Registry (RPDR) and patients for providing health information data.

