## Supplemental Methods for "Impact of Pulmonary and Sleep Disorders on COVID-19 Infection Severity in a Large Clinical Biobank"

Study sample

All patients provided written consent and were enrolled in the MGB Research Patient Data Registry EHR database, which contains date-stamped diagnosis, anthropometric, laboratory, procedure, and clinical note data [1,2]. We used a stringent combined data floor and loyalty cohort [3]  approach to minimize the number of patients with reduced documentation (and increased risk of false negative associations) in our open healthcare setting [4] by requiring at least three clinical notes, encounters, and diagnoses. We also required at least one diagnosis, encounter, laboratory result, medication, outpatient visit, routine visit [3] and vital sign obtained before 2020. Children (born after 1/1/2002), MGB employees, and patients living outside of Massachusetts or without available race/ethnicity, age, biological sex, or BMI values were excluded.

Data used

Structured data for diagnoses, procedures and laboratory results were obtained from the MGB RPDR EHR database, with additional Covid-specific data obtained from the MGB Covid-19 data mart and vaccination registries.

All patients were diagnosed with Covid-19, based on their first positive PCR test (we did not analyze Covid-19 reinfections). Majority variant date ranges were based on regional sequencing by the Broad Institute Covid-19 Genome Surveillance project (https://covid-19-sequencing.broadinstitute.org/). Transition dates from the pre-Delta to the Delta and Omicron variants were set at 7/3/2021 and 12/17/2021 respectively. Study collection ended at 6/1/2022. A breakthrough infection was defined as having occurred over 30 days after a second vaccination.

The date of first infection was used to set participant age, BMI target measurement date, and phases of infection. BMI was extracted from structured tables and from unstructured clinical notes using regular expressions. The two BMI measurements closest in time to the participant's defined age were averaged together to calculate the participant's defined BMI. The acute phase of infection for measuring ICU admissions, mechanical ventilation, and inpatient admissions was defined as -7 to +30 days relative to the PCR test. All diseases, procedures, and NLP terms were extracted at least 8 days prior to the PCR test. Laboratory measurements were extracted from -5 to -1 years prior to the PCR test.

Preexisting diseases were based on PheCode groupings of ICD9 and ICD10 codes [5,6] as detailed in **Table S1**. Our focus was on evaluating common and selected rare pulmonary disorders, common sleep disorders (with certain symptoms enriched in long Covid) [7–9]. Lymphangioleiomyomatosis and tuberous sclerosis complex were the targets of their respective ICD definitions, but other diseases could not be separated at the resolution of ICDs and PheCodes. Non-pulmonary/sleep diseases were also aggregated using PheCodes. Definitions for individual diseases are available at the PheCode website ( https://phewascatalog.org/phecodes and https://phewascatalog.org/phecodes_icd10cm ).

Unstructured clinical note concept terms were extracted from notes considering natural language processing (NLP) terms that mapped to Concept Unique Identifiers (CUIs) from the Unified Medical Language System using cTAKES [10,11]. Counts of each non-negated term were summed across all notes. Multimodal Automated Phenotyping (MAP) was used to improve case phenotyping by supplementing ICD codes with the count of their exact text matches located within clinical notes (*e.g.* count of "obstructive sleep apnea" phrases) [12]. MAP uses only a smaller number of CUIs that exactly match ICD diagnosis descriptions that are grouped within a single PheCode. We retained non-pulmonary/sleep MAP-defined diseases with a minimum 1% prevalence. Of 1,374 total MAP phenotypes, we retained 561. We also retained 9,839 of 75,259 non-negated NLP terms seen in ≥1% of patients with Covid-19 prior to the date of the PCR test.

1,647 procedures recorded in ≥1% of patients with Covid-19 were retained. Given the sparse nature of specific procedure counts, we also examined 171 procedures grouped as specified by the Agency for Healthcare Research and Quality Clinical Classification Software (CCS, https://www.hcup-us.ahrq.gov/toolssoftware/ccs10/ccs10.jsp). Procedure and CUI IDs for variables retained after the final LASSO regressions are listed in Table S6.

We documented evidence of continuous positive airway pressure (CPAP) use among patients with at least one clinical encounter and one clinical note from -365 to -8 days prior to the first positive PCR test. Total counts of five related procedures (CPT 94660, ICD9 93.90, ICD10 5A09357, and two internal codes) and the non-negative NLP term C0199451 were combined -365 to -8 days prior to the first positive PCR test. No other relevant CPAP NLP terms were present in our cTAKES-derived NLP data. CPAP evidence in the prior year was present for 2,946 / 4,027 (73.1%) of the eligible patients with sleep apnea.

In an exploratory aim, we examined whether complete blood cell count and other hematology laboratory results collected in patients between 1 – 5 years prior to infection were associated with Covid-19 severity. We also included measures of kidney function given the retention of kidney disease in the adaptive LASSO results described below. Results for the 23 laboratory measurements were available for 65 – 89% of the patients (Table S8). The median value of laboratory results in individual patients from across the 4-year period were tested in logistic regression with and without rank normalization.

Statistical analyses

Logistic regression included the covariates age, sex at birth, obesity classification, race/ethnicity, and breakthrough infection status. The effect of CPAP evidence was tested using logistic regression with further adjustment for a single CPAP evidence term.

Adaptive LASSO regression used and 80% training and 20% testing sets, each with 20-fold cross-validation. Non-zero coefficient terms from the training runs were included in the testing runs. Non-zero coefficient terms from the testing runs were included in the final analyses of combined training and testing participants. The covariates age, sex, BMI, race/ethnicity, breakthrough infection status, total healthcare encounters, and days into the pandemic were forced into all training, testing and combined sample runs. Given reduced sample size and non-random ascertainment (which may correlate with multimorbidity), the laboratory values were not included in the LASSO analyses. Nonzero coefficient adaptive LASSO terms were identified in separate analyses of diseases, procedures, CCS procedures, and NLP terms in combined-sex and sex-stratified analyses. We opted to include sex-specific terms, which could alternatively be due to true associations or ascertainment biases, in both combined-sex analyses and sex-stratified analyses. We tested the association of individual LASSO terms and Covid-19 using logistic regression, adjusting for age, sex at birth, obesity classification, race/ethnicity, and breakthrough infection status. Finally, the potential effect of lead LASSO variables on disease associations with Covid-19 infection severity was assessed using logistic regression with standard covariates that included and excluded individual LASSO variables as covariates. The sample size for logistic regression calculations considering laboratory tests was reduced to participants who received the laboratory test.

2 Karlson EW, Boutin NT, Hoffnagle AG, *et al.* Building the Partners HealthCare Biobank at Partners Personalized Medicine: Informed Consent, Return of Research Results, Recruitment Lessons and

Operational Considerations. *J Med* 2016;**6**.

3 Weber GM, Adams WG, Bernstam EV, *et al.* Biases introduced by filtering electronic health records for patients with “complete data.” *J Am Med Inform Assoc JAMIA* 2017;**24**:1134–41. doi:10.1093/jamia/ocx071

4 Robinson JR, Wei W-Q, Roden DM, *et al.* Defining Phenotypes from Clinical Data to Drive Genomic Research. *Annu Rev Biomed Data Sci* 2018;**1**:69–92. doi:10.1146/annurev-biodatasci-080917-013335

5 Wei W-Q, Bastarache LA, Carroll RJ, *et al.* Evaluating phecodes, clinical classification software, and ICD-9-CM codes for phenome-wide association studies in the electronic health record. *PloS One* 2017;**12**:e0175508. doi:10.1371/journal.pone.0175508

6 Wu P, Gifford A, Meng X, *et al.* Mapping ICD-10 and ICD-10-CM Codes to Phecodes: Workflow Development and Initial Evaluation. *JMIR Med Inform* 2019;**7**:e14325. doi:10.2196/14325

7 Cade BE, Dashti HS, Hassan SM, *et al.* Sleep Apnea and COVID-19 Mortality and Hospitalization. *Am J Respir Crit Care Med* 2020;**202**:1462–4. doi:10.1164/rccm.202006-2252LE

8 Jones SE, Maisha FI, Strausz SJ, *et al.* The public health impact of poor sleep on severe COVID-19, influenza and upper respiratory infections. *MedRxiv Prepr Serv Health Sci* 2022;:2022.02.16.22271055. doi:10.1101/2022.02.16.22271055

9 Davis HE, Assaf GS, McCorkell L, *et al.* Characterizing long COVID in an international cohort: 7 months of symptoms and their impact. *EClinicalMedicine* 2021;**38**:101019. doi:10.1016/j.eclinm.2021.101019

10 Savova GK, Masanz JJ, Ogren PV, *et al.* Mayo clinical Text Analysis and Knowledge Extraction System (cTAKES): architecture, component evaluation and applications. *J Am Med Inform Assoc JAMIA* 2010;**17**:507–13. doi:10.1136/jamia.2009.001560

11 Bodenreider O. The Unified Medical Language System (UMLS): integrating biomedical terminology. *Nucleic Acids Res* 2004;**32**:D267-270. doi:10.1093/nar/gkh061

12 Liao KP, Sun J, Cai TA, *et al.* High-throughput multimodal automated phenotyping (MAP) with application to PheWAS. *J Am Med Inform Assoc* Published Online First: 7 August 2019. doi:10.1093/jamia/ocz066
