## Supplemental Figures for "Impact of Pulmonary and Sleep Disorders on COVID-19 Infection Severity in a Large Clinical Biobank"

Figure S1. Combined sex death outcome LASSO term covariate adjustments −log10(p) differences

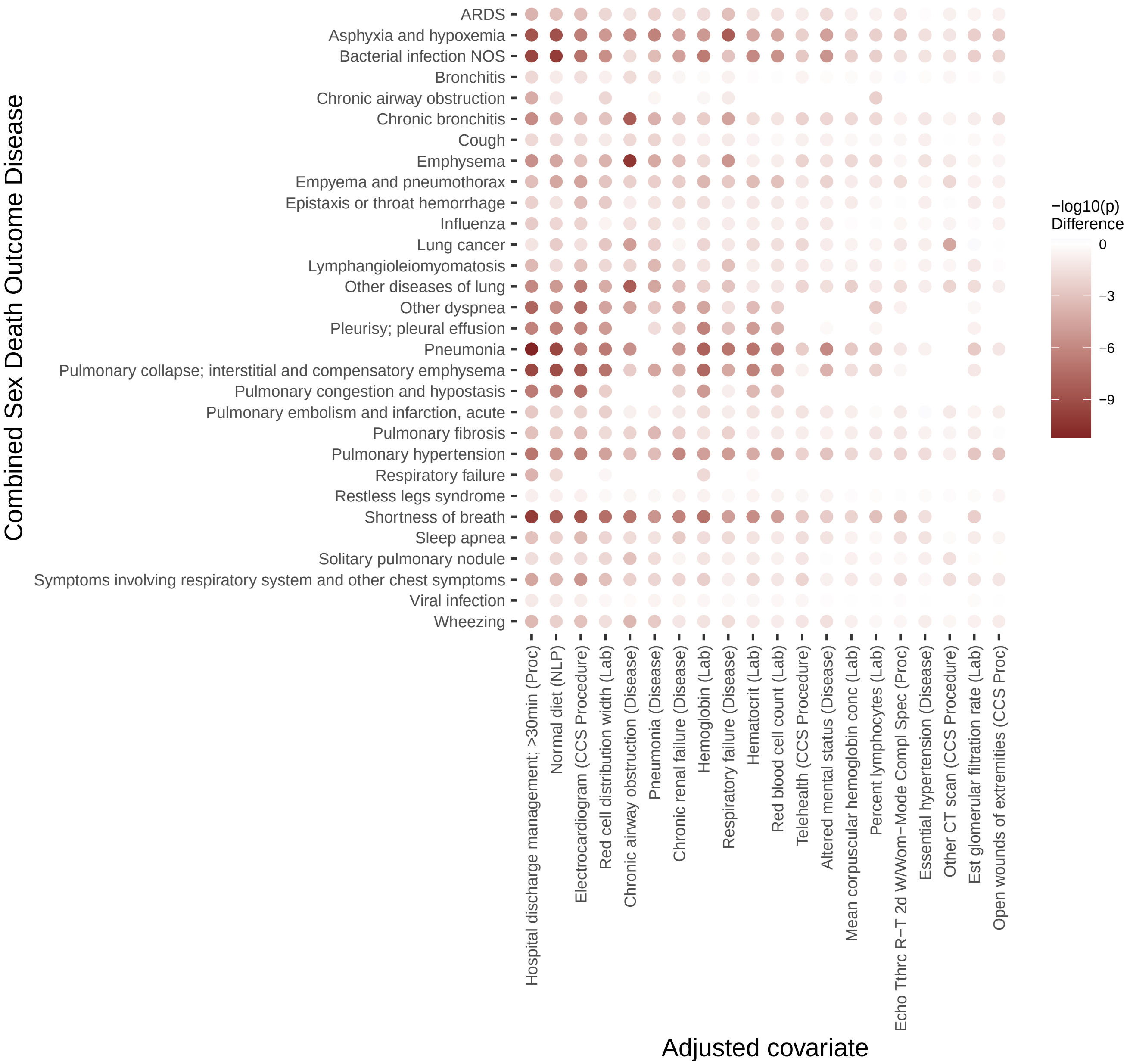

Figure S2. Combined sex composite outcome LASSO term covariate adjustments

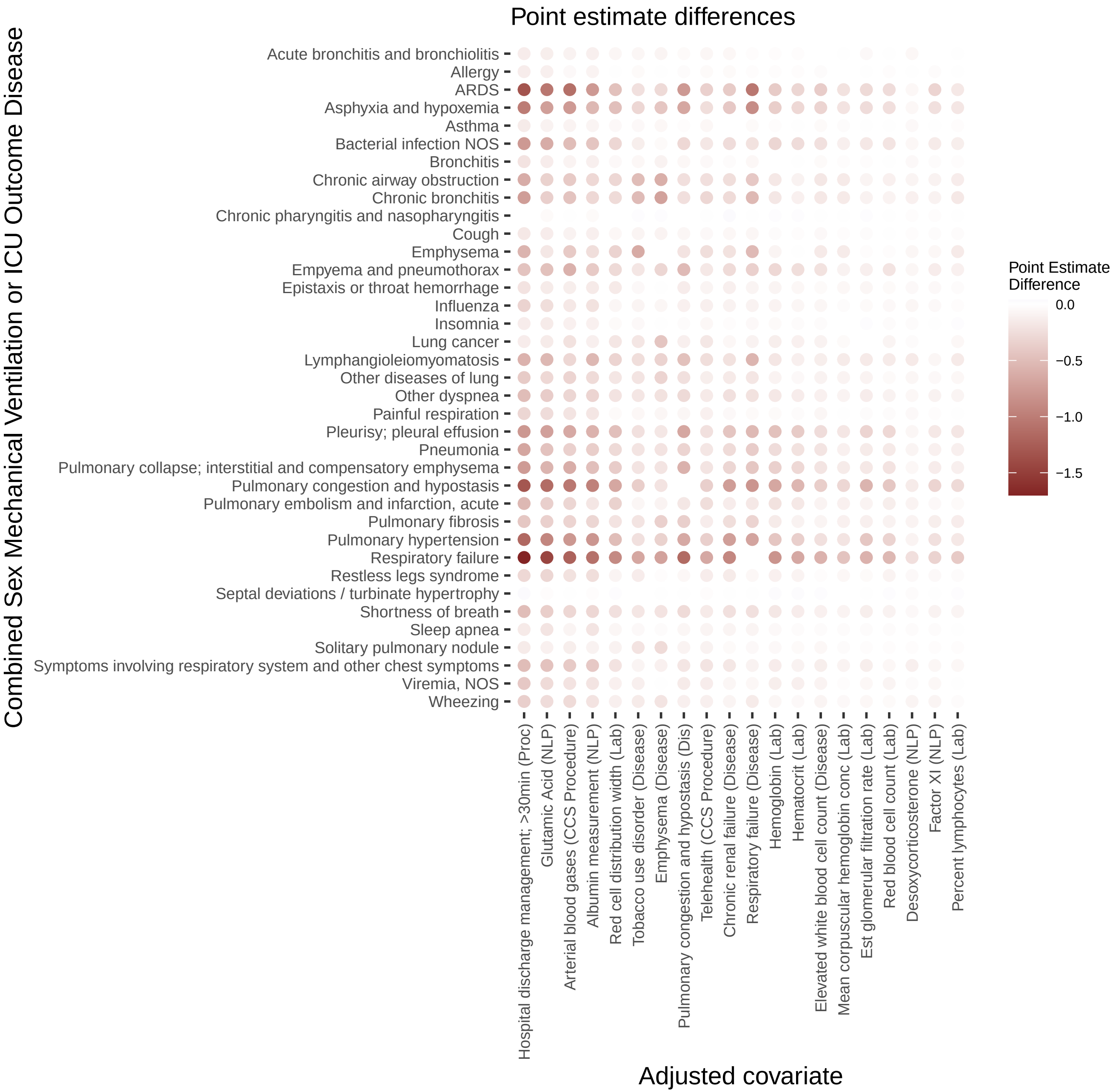

### Figure S3. Combined sex composite outcome LASSO term covariate adjustments −log10(p) differences

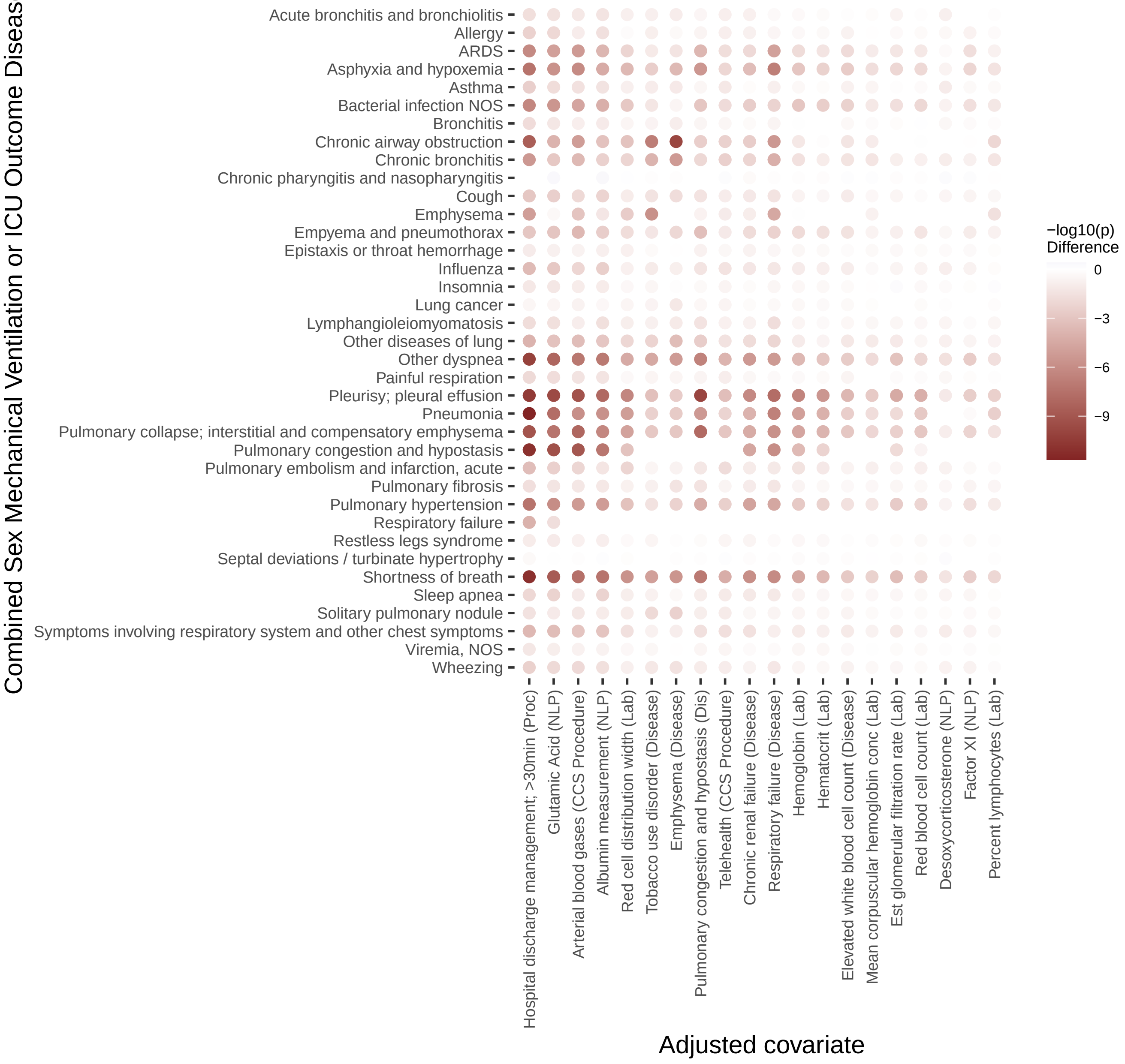

Figure S4. Combined sex inpatient outcome LASSO term covariate adjustments

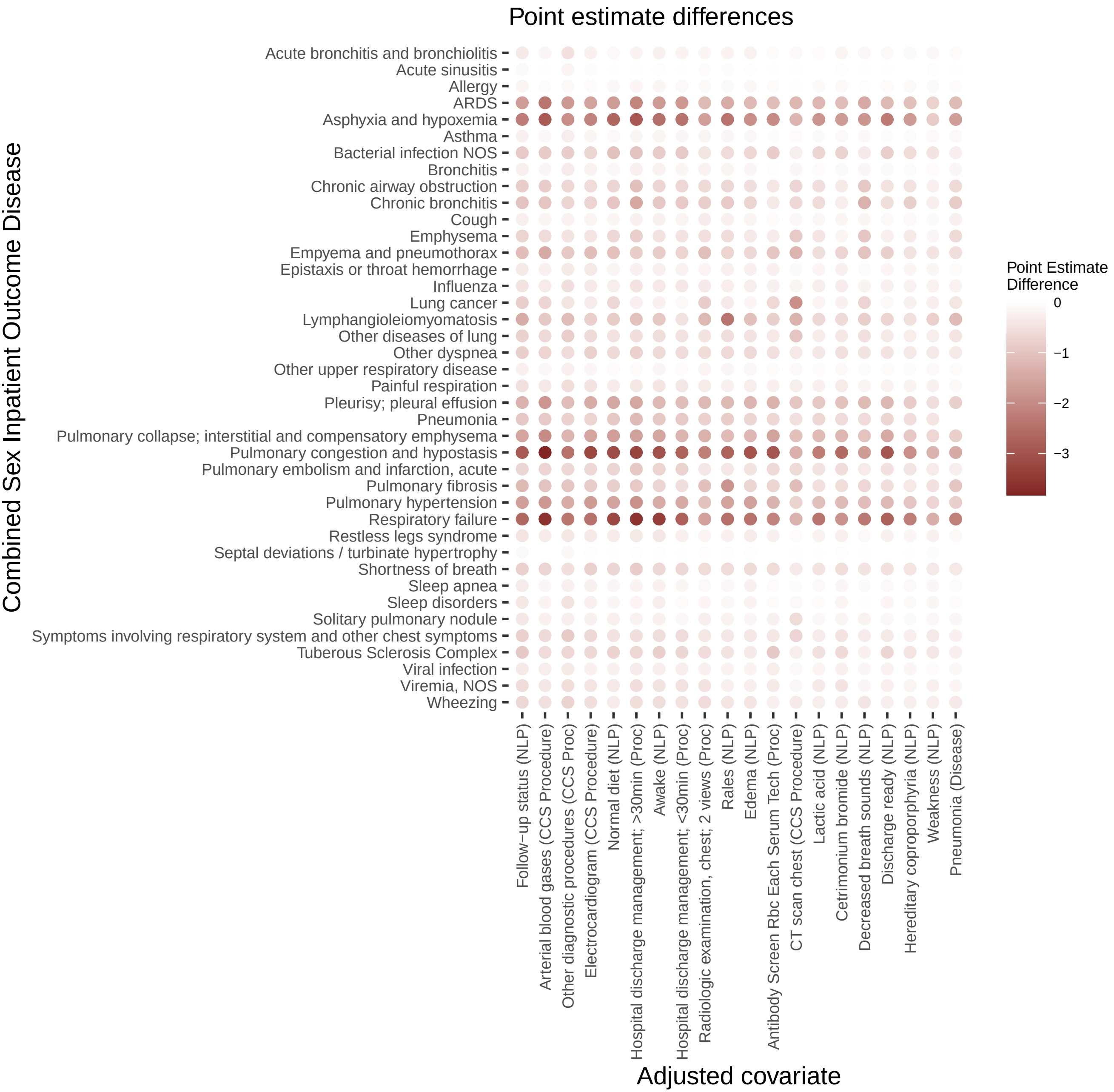

Figure S5. Combined sex inpatient outcome LASSO term covariate adjustments
−log10(p) differences

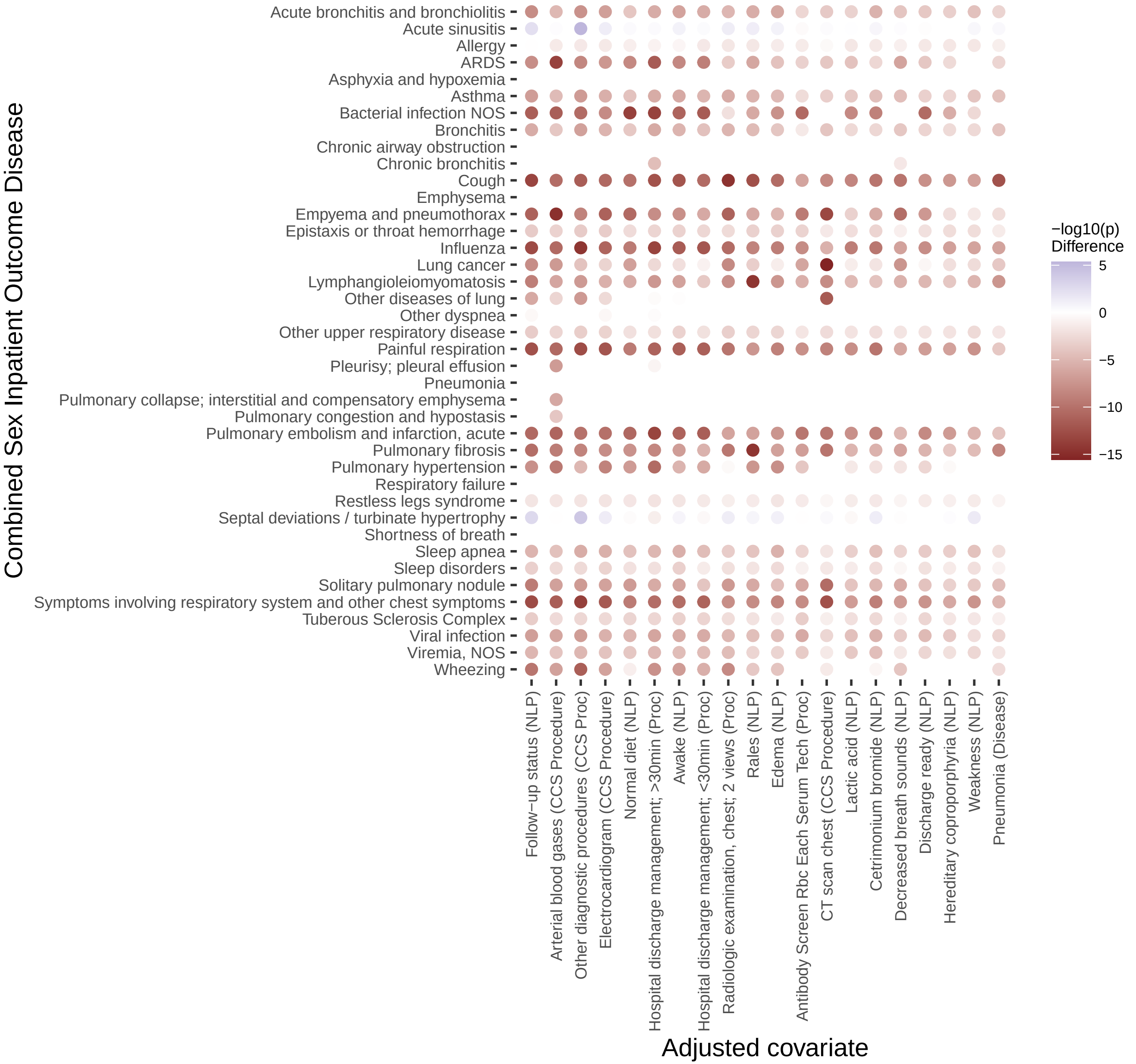

Figure S6. Men death outcome LASSO term covariate adjustments

Point estimate differences

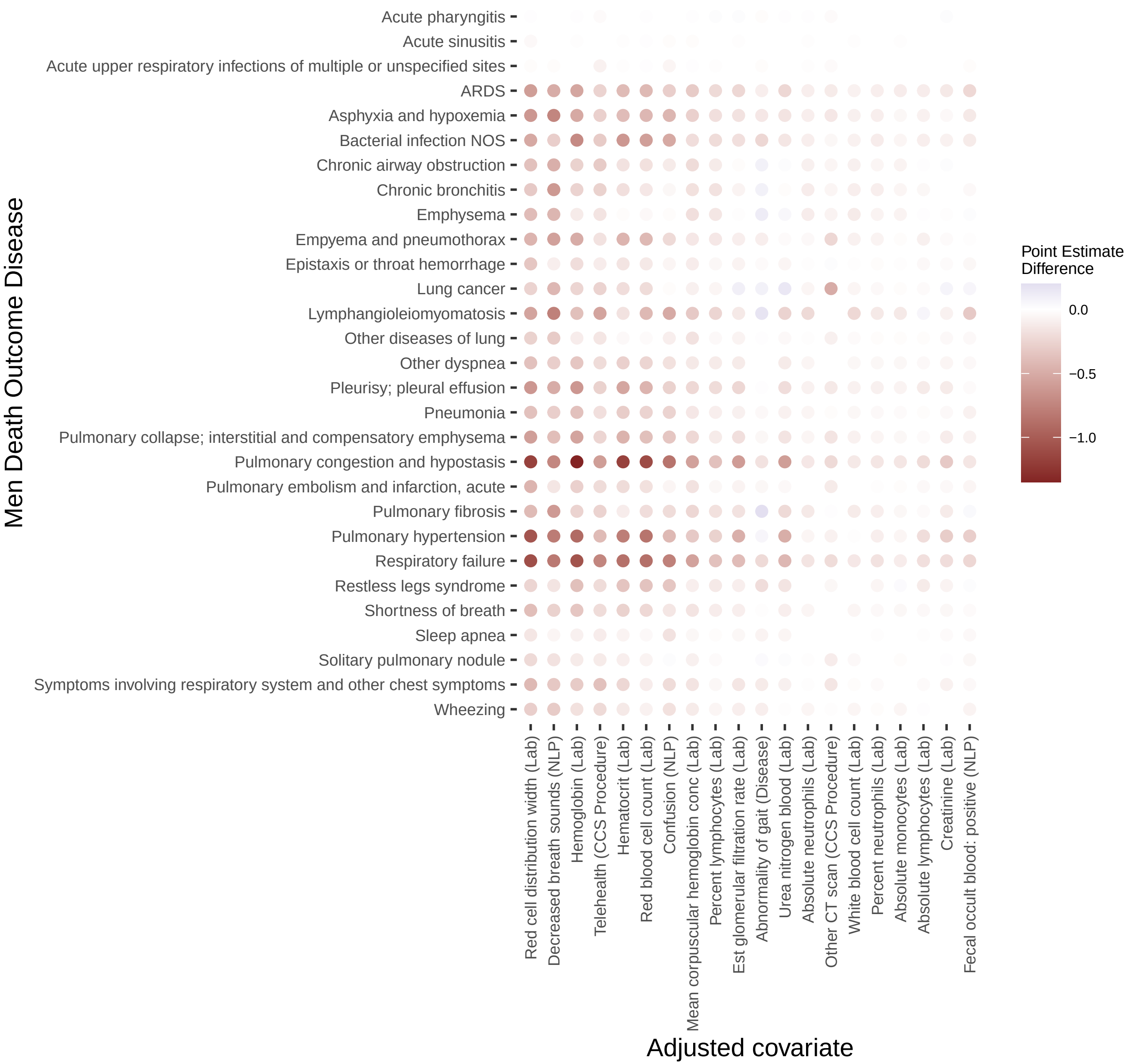

Figure S7. Men death outcome LASSO term covariate adjustments −log10(p) differences

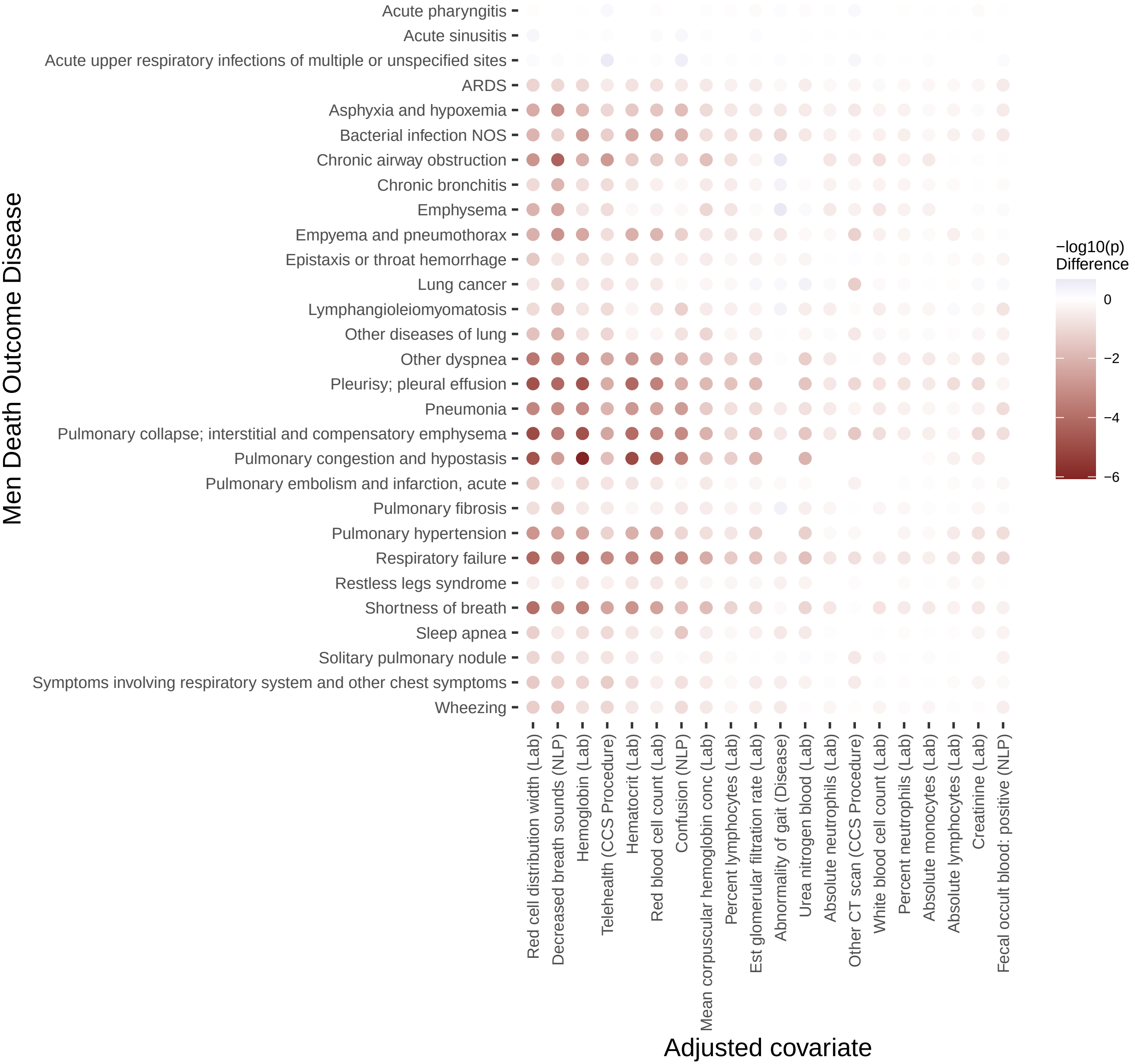

### Figure S8. Men composite outcome LASSO term covariate adjustments

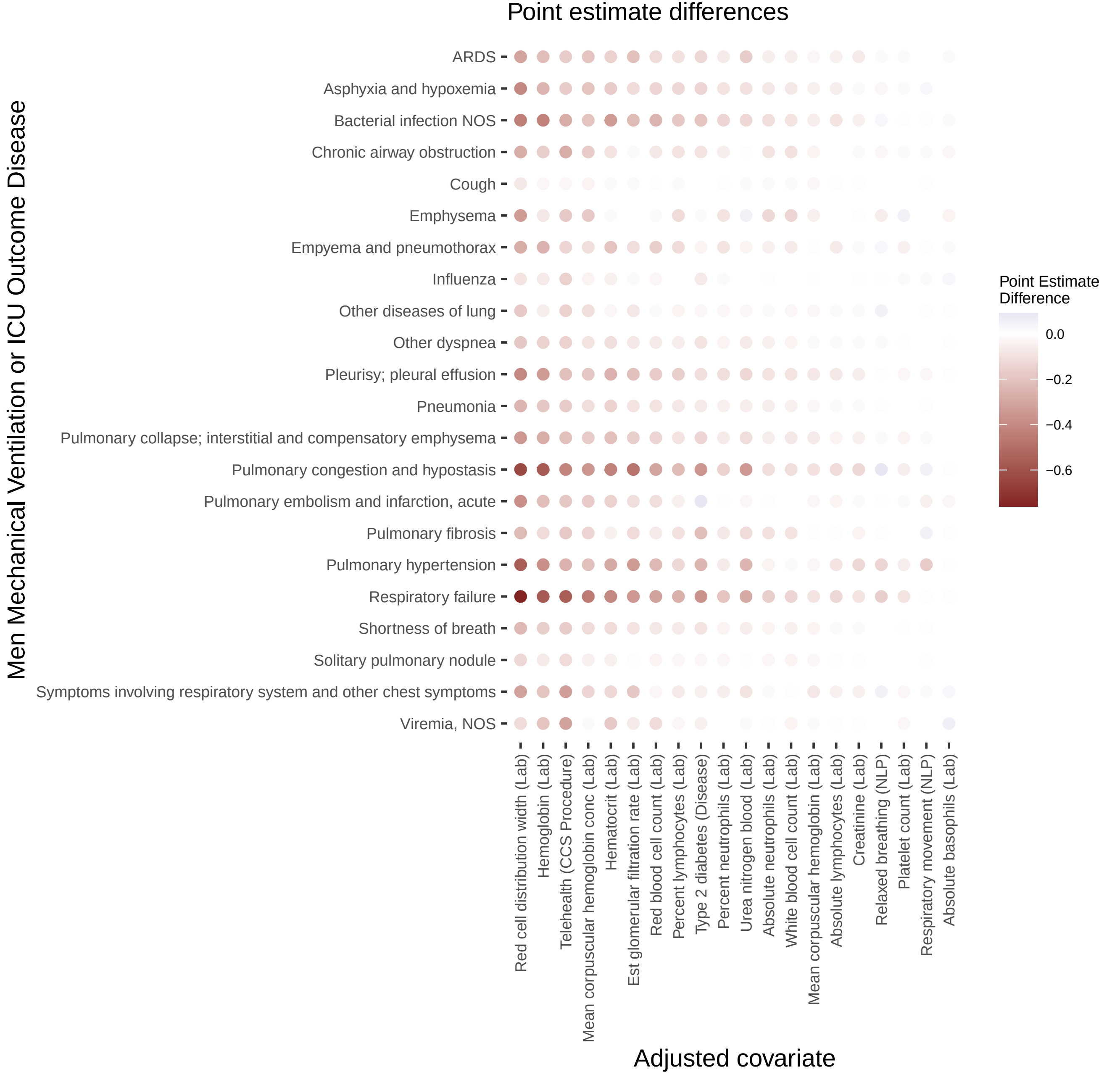

### Figure S9. Men composite outcome LASSO term covariate adjustments −log10(p) differences

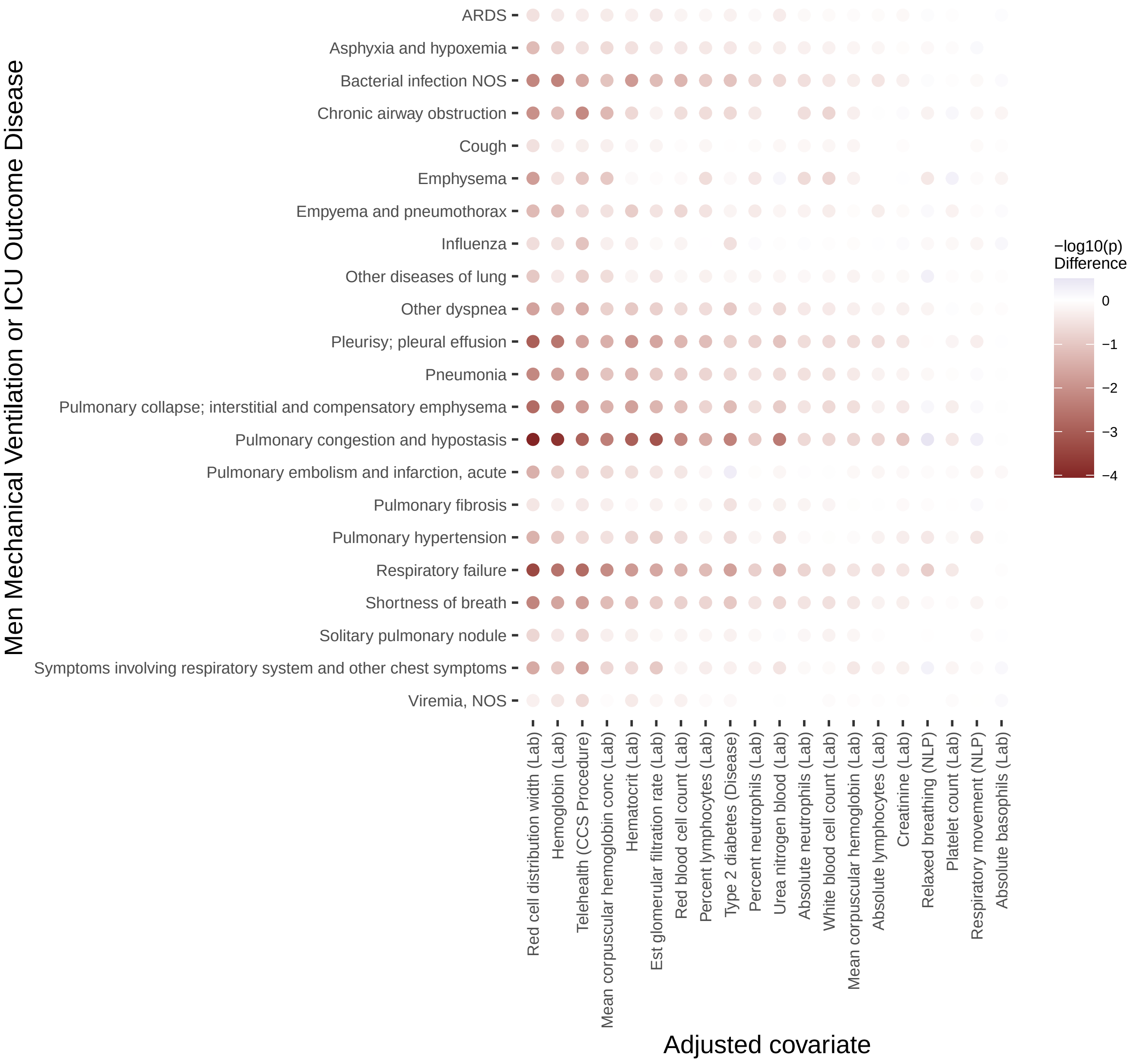

### Figure S10. Men inpatient outcome LASSO term covariate adjustments

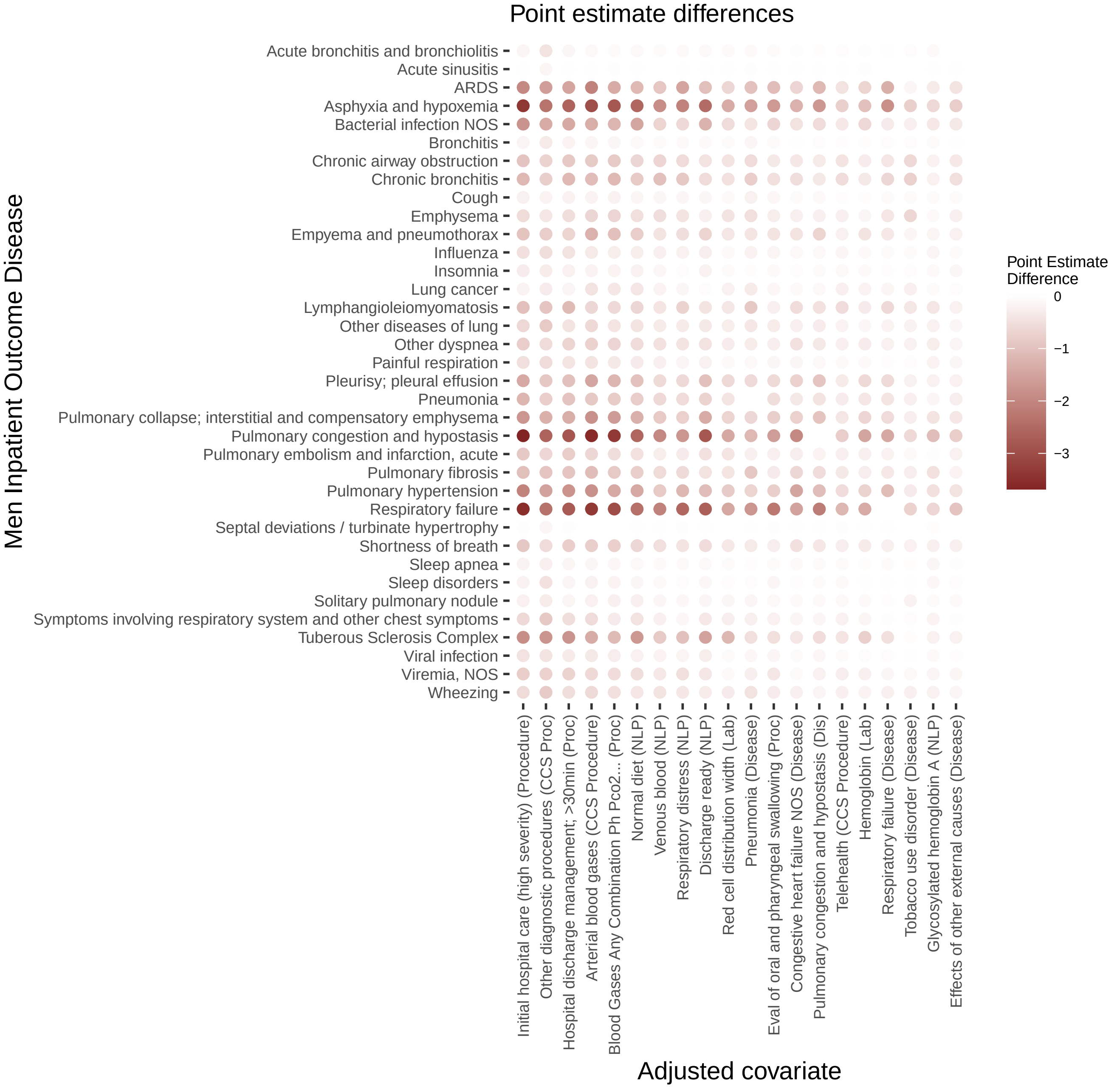

Figure S11. Men inpatient outcome LASSO term covariate adjustments −log10(p) differences

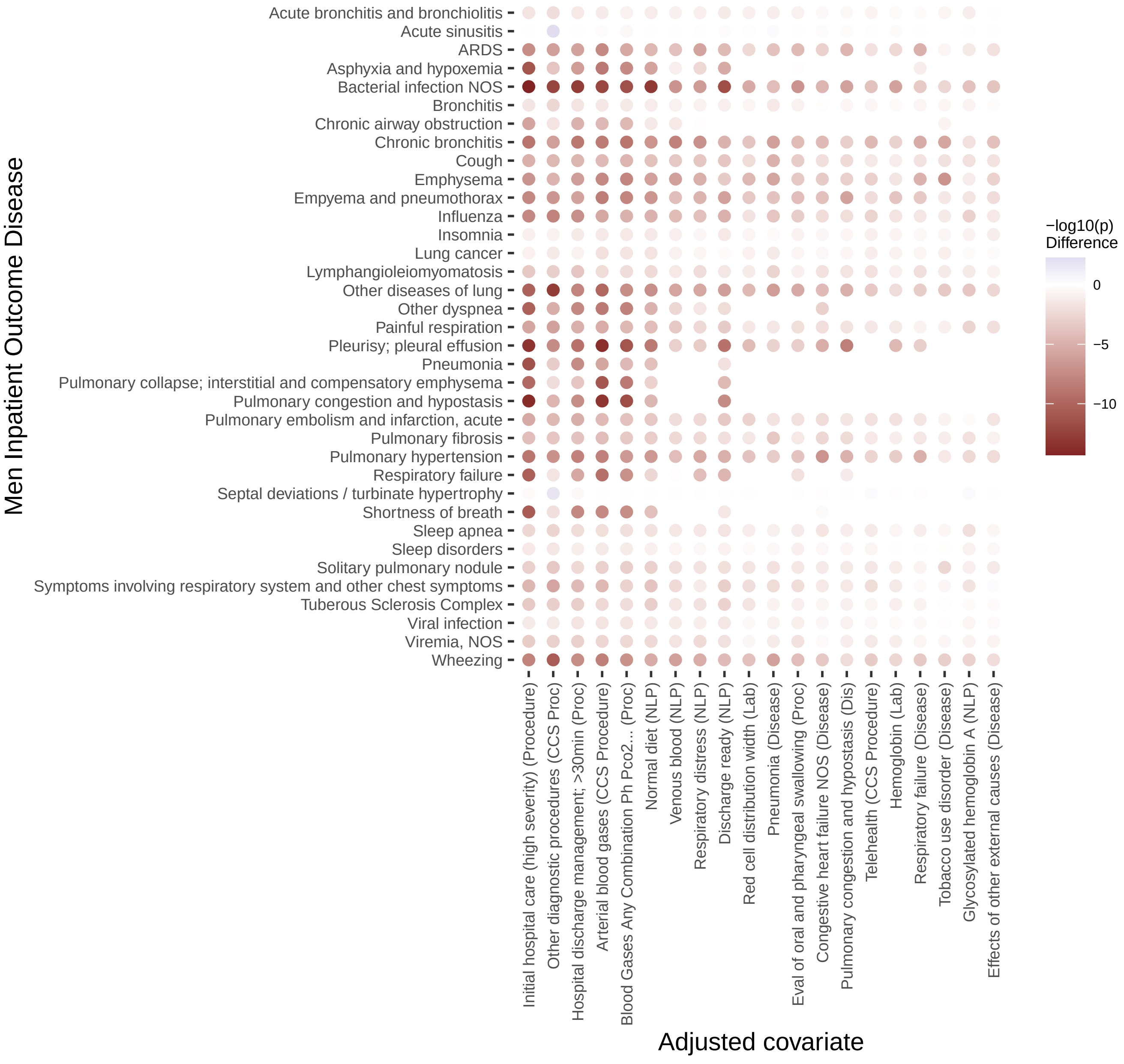

Figure S12. Women death outcome LASSO term covariate adjustments

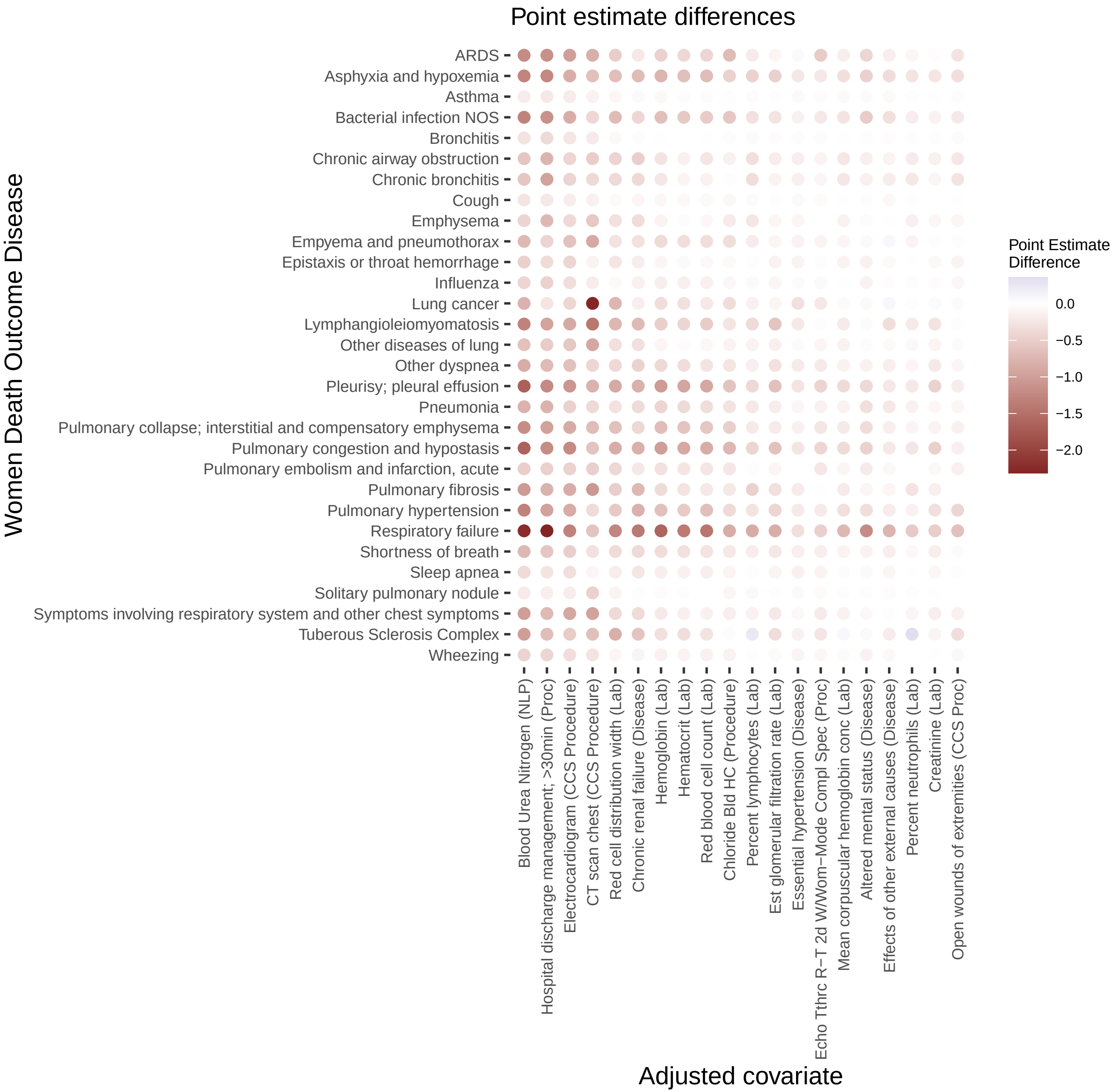

Figure S13. Women death outcome LASSO term covariate adjustments −log10(p) differences

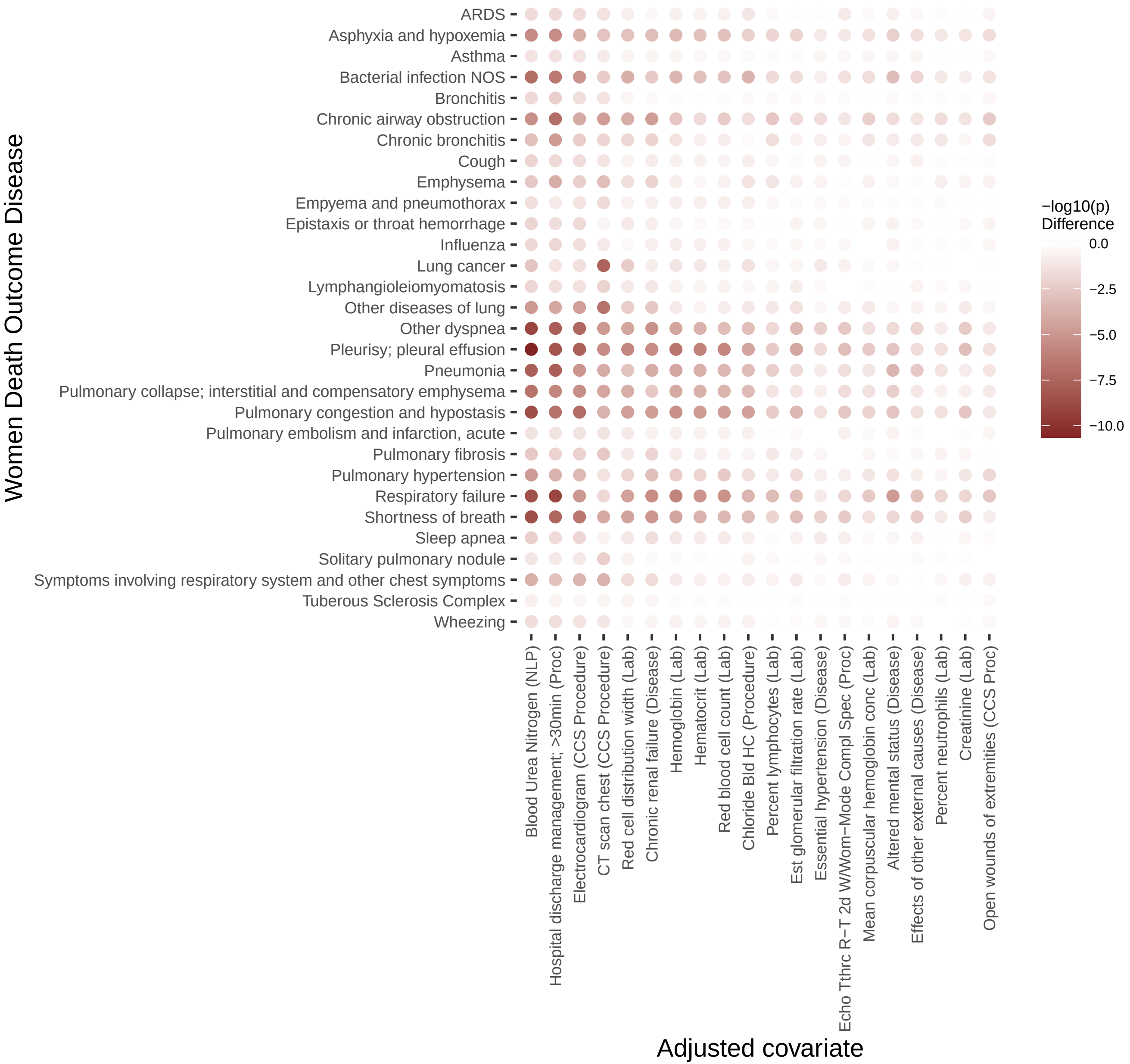

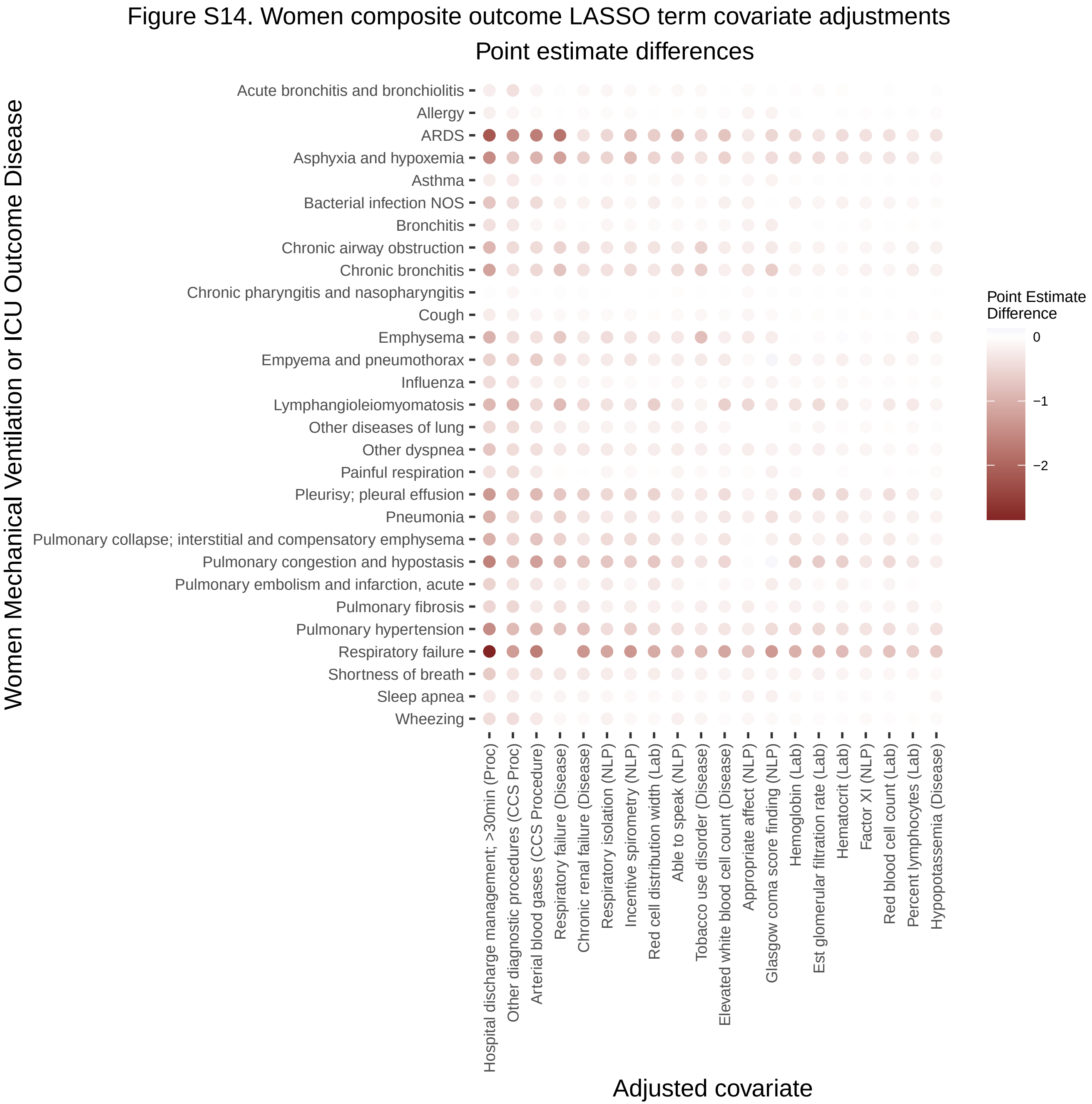

Figure S14. Women composite outcome LASSO term covariate adjustments
Point estimate differences
Women Mechanical Ventilation or ICU Outcome Disease
Acute bronchitis and bronchiolitis
Allergy
ARDS
Asphyxia and hypoxemia
Asthma
Bacterial infection NOS
Bronchitis
Chronic airway obstruction
Chronic bronchitis
Chronic pharyngitis and nasopharyngitis
Cough
Emphysema
Empyema and pneumothorax
Influenza
Lymphangioleiomyomatosis
Other diseases of lung
Other dyspnea
Painful respiration
Pleurisy; pleural effusion
Pneumonia
Pulmonary collapse; interstitial and compensatory emphysema
Pulmonary congestion and hypostasis
Pulmonary embolism and infarction, acute
Pulmonary fibrosis
Pulmonary hypertension
Respiratory failure
Shortness of breath
Sleep apnea
Wheezing
Hospital discharge management; >30min (Proc)
Other diagnostic procedures (CCS Proc)
Arterial blood gases (CCS Procedure)
Respiratory failure (Disease)
Chronic renal failure (Disease)
Respiratory isolation (NLP)
Incentive spirometry (NLP)
Red cell distribution width (Lab)
Able to speak (NLP)
Tobacco use disorder (Disease)
Elevated white blood cell count (Disease)
Appropriate affect (NLP)
Glasgow coma score finding (NLP)
Hemoglobin (Lab)
Est glomerular filtration rate (Lab)
Hematocrit (Lab)
Factor XI (NLP)
Red blood cell count (Lab)
Percent lymphocytes (Lab)
Hypopotassemia (Disease)
Adjusted covariate
Point Estimate Difference
0
−1
−2

Figure S15. Women composite outcome LASSO term covariate adjustments −log10(p) differences

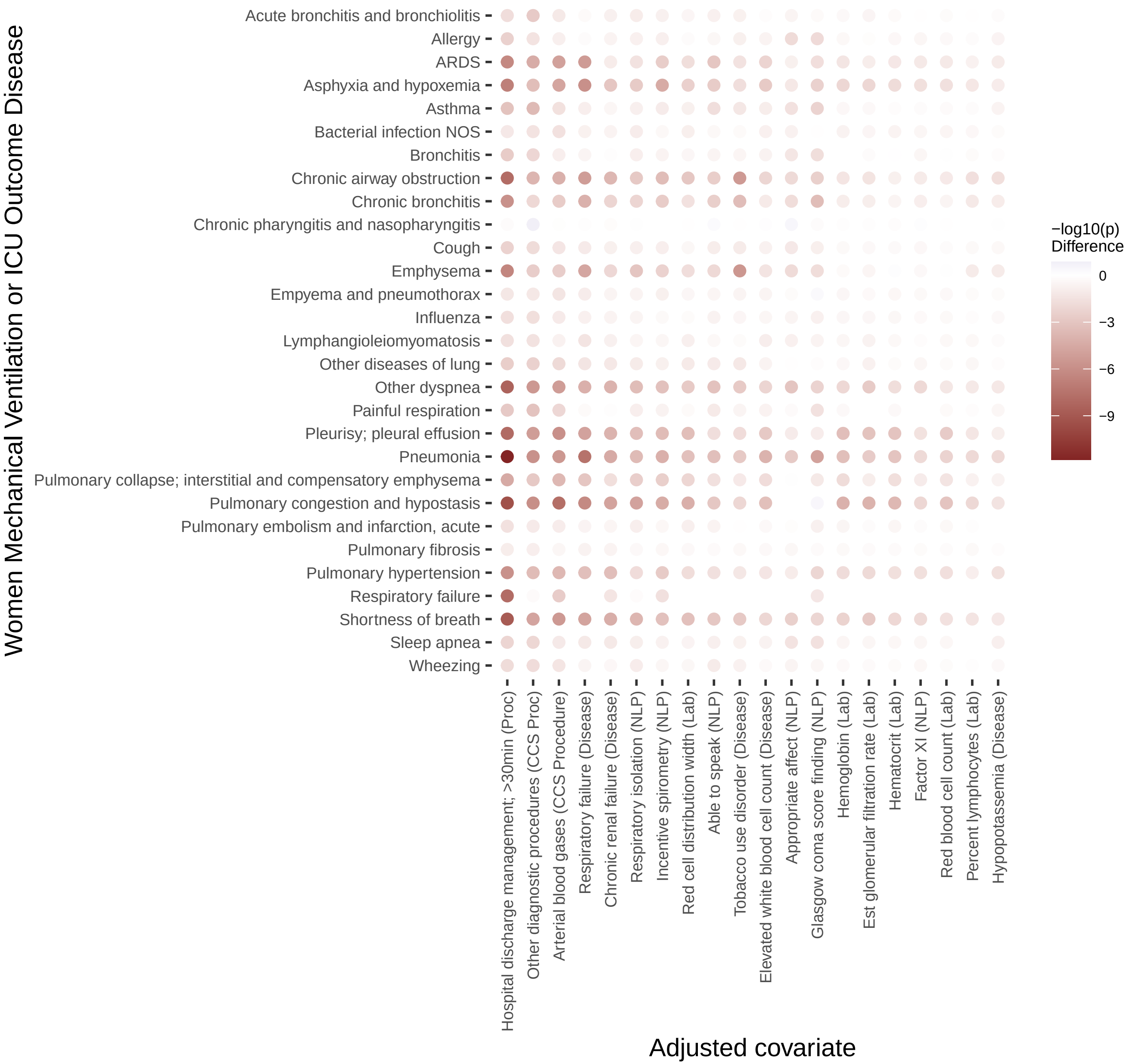

Figure S16. Women inpatient outcome LASSO term covariate adjustments

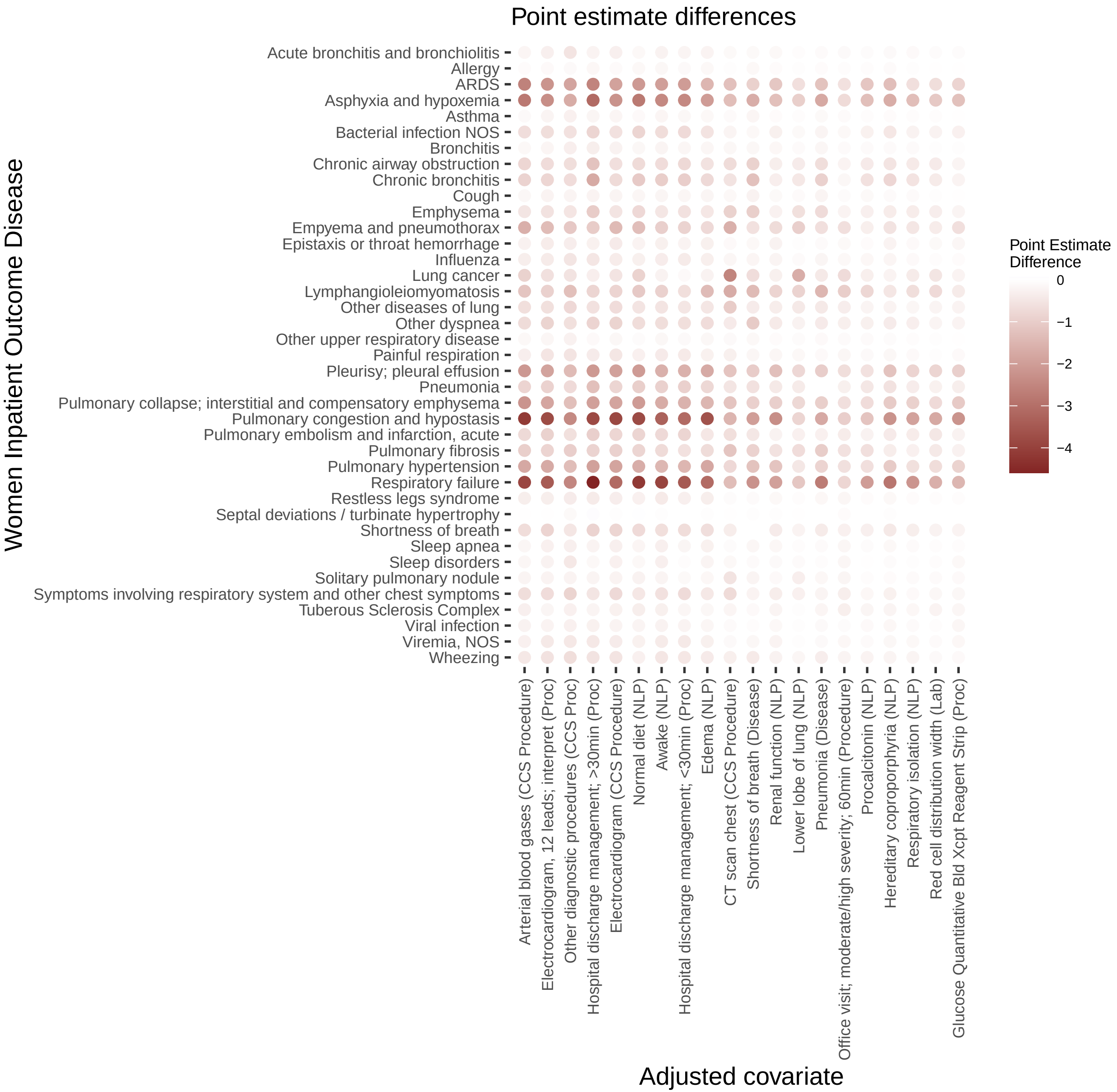

Figure S16. Women inpatient outcome LASSO term covariate adjustments −log10(p) differences

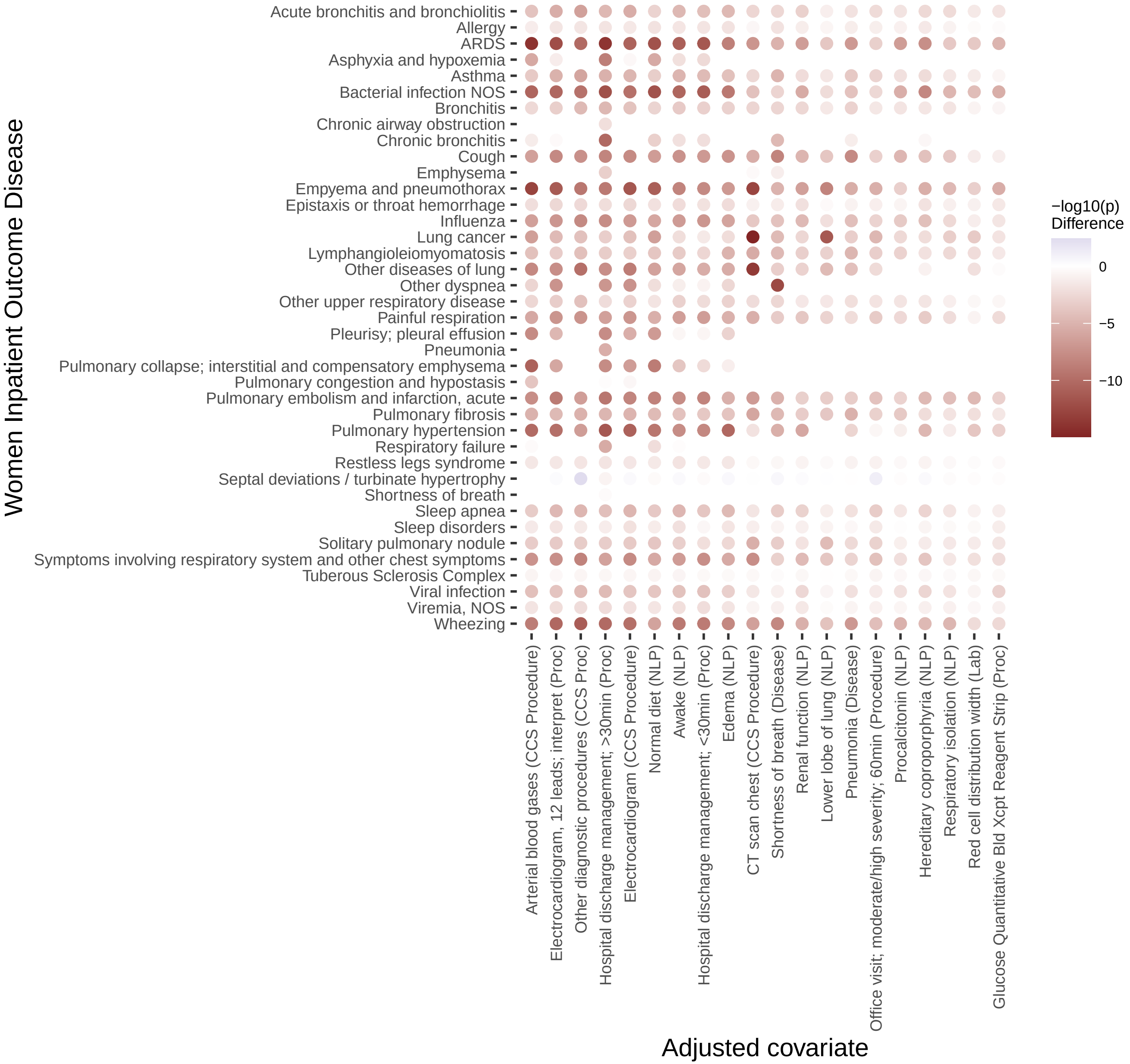
